# Assessing the Role of Model Complexity in Virtual Clinical Trial Outcomes

**DOI:** 10.64898/2025.12.22.25342808

**Authors:** Jana L. Gevertz, Joanna R. Wares

## Abstract

Virtual clinical trials (VCTs) hold significant promise for improving the drug development process, yet their predictive reliability depends critically on design decisions that remain poorly understood. This study examines how model complexity influences VCT outcomes, as well as how the choice of prior parameter distributions and virtual patient inclusion criteria affects those outcomes. Using oncolytic virotherapy treatment of murine tumors as a case study, we compared a relative hierarchy of three mathematical models of increasing complexity under different parameter priors (uniform and normal distributions) and two inclusion methods (accept-or-reject and accept-or-perturb). Our results demonstrate that the simplest model produces a plausible population that inadequately spans the feasible trajectory space, potentially missing critical inter-patient heterogeneity. However, we found diminishing returns beyond intermediate model complexity, as both the intermediate and complex models captured similar ranges of patient responses across dosing protocols. Notably, the accept-or-reject method generated posterior parameter distributions that are more likely to resemble the chosen prior, possibly overly reducing inter-patient variability in treatment responses, particularly at high doses. In contrast, the accept-or-perturb inclusion criteria produced more robust results that were less sensitive to prior assumptions. These findings suggest that VCT design should prioritize models with sufficient biological detail to capture key mechanisms without unnecessary complexity, paired with inclusion criteria that avoid over-constraining plausible populations to match potentially unrealistic prior assumptions.

## 1 Introduction

Virtual clinical trials (VCTs), or in silico trials, offer a powerful framework for testing therapies by complementing traditional clinical studies with computational assessments of efficacy, safety, and trial design. Unlike standard clinical trials, which test interventions directly on patients, VCTs utilize virtual patient populations and mathematical models [1–5].

Conducting a virtual clinical trial is a multi-step process [5, 6] that begins with identifying an intervention of interest. From there, a fit-for-purpose mechanistic model is developed, calibrated to available data, and ideally validated against untrained data. The next step of the process is determining the simulated patients that get “enrolled” in the virtual clinical trial. First, a plausible population is created consisting of model parameterizations that satisfy all imposed biological and physiological constraints [6]. If population-level clinical data is available, this plausible population can be statistically weighted and filtered to generate a true virtual population that matches the baseline characteristics and joint distributions of a specific clinical cohort (see, for instance [7]). The virtual clinical trial is then conducted on either the plausible or virtual population, allowing for the prediction of both population-level and subgroup-level treatment responses [6].

Despite the growing importance of VCTs across a range of clinical indications, including oncology [8–12], metabolic diseases [13–15], immunology [16], and more, a number of open questions remain about the design of these trials. One important question is how to determine which simulated patients get enrolled in a virtual clinical trial. Just as the lack of diversity in clinical trial populations can result in clinical recommendations that are either less effective, or potentially even harmful for certain subpopulations [17], the heterogeneity captured in a plausible population and subsequent virtual patient population can significantly influence the recommendations made from a virtual clinical trial.

In a previous work [18], we quantified the impact of two VCT design choices: the selection of the prior distribution for each parameter that varies across plausible patients and the criteria used to determine if a parameterization sampled from the priors gets included or excluded in the plausible population. We found that the heterogeneity of the posterior distributions in the plausible population was more affected by which prior distribution was used, rather than the choice of inclusion/exclusion criteria. However we found that the percent of treatment responders in the plausible population was more sensitive to the inclusion/exclusion criteria utilized. Recent work in this area by Schirru and colleagues [19] rigorously explored the ability of different patient generation techniques to capture the diversity of parameter combinations that reflect biological variability.

In this work, we tackle a different question about VCT design. We ask how the complexity of the mathematical model used in the trial affects the heterogeneity of the plausible populations and the trial outcomes. We will explore this question using a previously studied and modeled preclinical system of oncolytic viruses in a murine melanoma model [20, 21]. Oncolytic virotherapy is a cancer treatment that exposes tumors to viruses, called oncolytic viruses (OVs), that are engineered to specifically replicate in cancer cells but not healthy cells [22–24]. OV replication within a cancer cell eventually causes the infected cancer cell to lyse. This bursting has two effects, the first being that the infected cell is killed and releases viruses into the tissue space which can infect other cancer cells. Second, this lysis releases tumor antigens and inflammatory signals which can help recruit tumor-targeting immune cells into the tumor microenvironment [22–25].

Researchers have developed a wide range of mathematical models to investigate the dynamics of oncolytic virotherapy. From a biological perspective, these models can vary greatly in their complexity. Earlier models focus only on viral lysis [26–29], whereas others aggregate the immune effects into a single term [30–32] and yet others aim to describe more granular details of the immune response [21, 33, 34]. Models can also vary in their mathematical complexity, from ordinary differential equation models that describe well-mixed populations to agent-based models that explicitly model the spatial distribution of cancer cells and OVs [35–37].

Model-based VCTs have been used to better understand the mechanisms driving inter-patient response heterogeneity to oncolytic virotherapy. Cassidy et al. [38] integrated a model of tumor-immune dynamics into a VCT platform to evaluate combination protocols of GM-CSF immunotherapy and talimogene laherparepvec (T-VEC) virotherapy in virtual patient cohorts with late-stage melanoma. The VCT identified a clinically-actionable combination schedule that significantly improved virtual patient outcomes when compared to GM-CSF and T-VEC monotherapies, and a standard combination strategy [38]. In another study, Jenner and colleagues [3] conducted a VCT to optimize combination protocols involving sequential administrations of an enhancer virus (oncolytic vaccinia) and a heterologous primary virus (vesicular stomatitis virus). They found that the optimal injection lag between doses sensitively depends on how aggressively a patient’s tumor is growing [3]. These studies demonstrate that VCTs can effectively stratify patients and optimize dosing schedules to maximize oncolytic virotherapy efficacy.

The goal of the current work is to study how model complexity influences the heterogeneity of plausible populations and the outcomes of a virtual clinical trial, using oncolytic virotherapy as a test case. This manuscript is organized as follows. In the Methods section, we introduce a previously-published experimental dataset describing murine melanoma dynamics in response to oncolytic virotherapy. We next present three ordinary differential equation models, each progressively more complex in terms of the number of variables and parameters, that well capture the average response to OV therapy. We also review two commonly-used methods for generating plausible populations from a mathematical model and explain the virtual clinical trial we will be conducting in this study. In the Results & Discussion section, we explore how model complexity influences the outcomes of the virtual clinical trial. We particularly focus on the diversity of patients in the plausible population and the heterogeneity in treatment response across a range of OV doses. We also begin to explore the question of whether features associated with a specific treatment response are consistent across models, or depend on model complexity. We conclude with a discussion of what the results tell us about the complexity of the models we use to conduct virtual clinical trials.

## 2 Methods

The foundation of a virtual clinical trial is a data-informed mathematical model. However, there is no simple strategy for identifying the “best” model for a dataset. Information criteria (IC) can be used to identify the most parsimonious model that best balances goodness-of-fit and model complexity (measured through the number of parameters), subject to some assumptions about the underlying noise structure of the data [39]. Oftentimes, several models have comparable IC values, confounding the selection of the most parsimonious model [40–43].

IC values also do not assess whether individual parameters are practically identifiable, meaning they can be uniquely determined from available data [44]. It is well-established that more complex models inevitably contain many non-identifiable parameters, though the consequences of this lack of identifiability is nuanced from the perspective of conducting a VCT [45]. If indeed diverse parametric combinations can yield the same biological outcome, a model with all identifiable parameters would generate virtual populations that are restrictively homogeneous. However, a model with many non-identifiable parameters runs the risk of making nonsensical predictions when extrapolating to scenarios far from the calibration data [45].

Thus, while both IC and identifiability analysis are essential tools for model selection, neither can provide the “correct” model to use for a VCT. Further, while both of these methods seek to establish the “right” level of model complexity based on available data, it is ultimately the question of interest that dictates the level of detail necessary to include [46–48]. For these reasons, in this work, we consider a relative hierarchy of three ordinary differential equations models of increasing complexity in terms of the number of state variables and model parameters. Using these models, we systematically study how model complexity influences the outcome of a virtual clinical trial.

### 2.1 Data

We consider previously-published experimental data from a preclinical murine trial describing the treatment of melanoma using an engineered oncolytic adenovirus, Ad-ΔB7 [20]. In this study, 5*×*10^5^ B16-F10 melanoma cells were subcutaneously implanted into male C57BL/6 mice. The available data consists of tumor volume measurements over a 16-day period, both in the absence of treatment and subject to an intratumoral dose of 10^10^ virions on days 0, 2, and 4 [20]. The individual time-course data for the OV-treated mice are shown in Figure 1A.

**Fig. 1.**
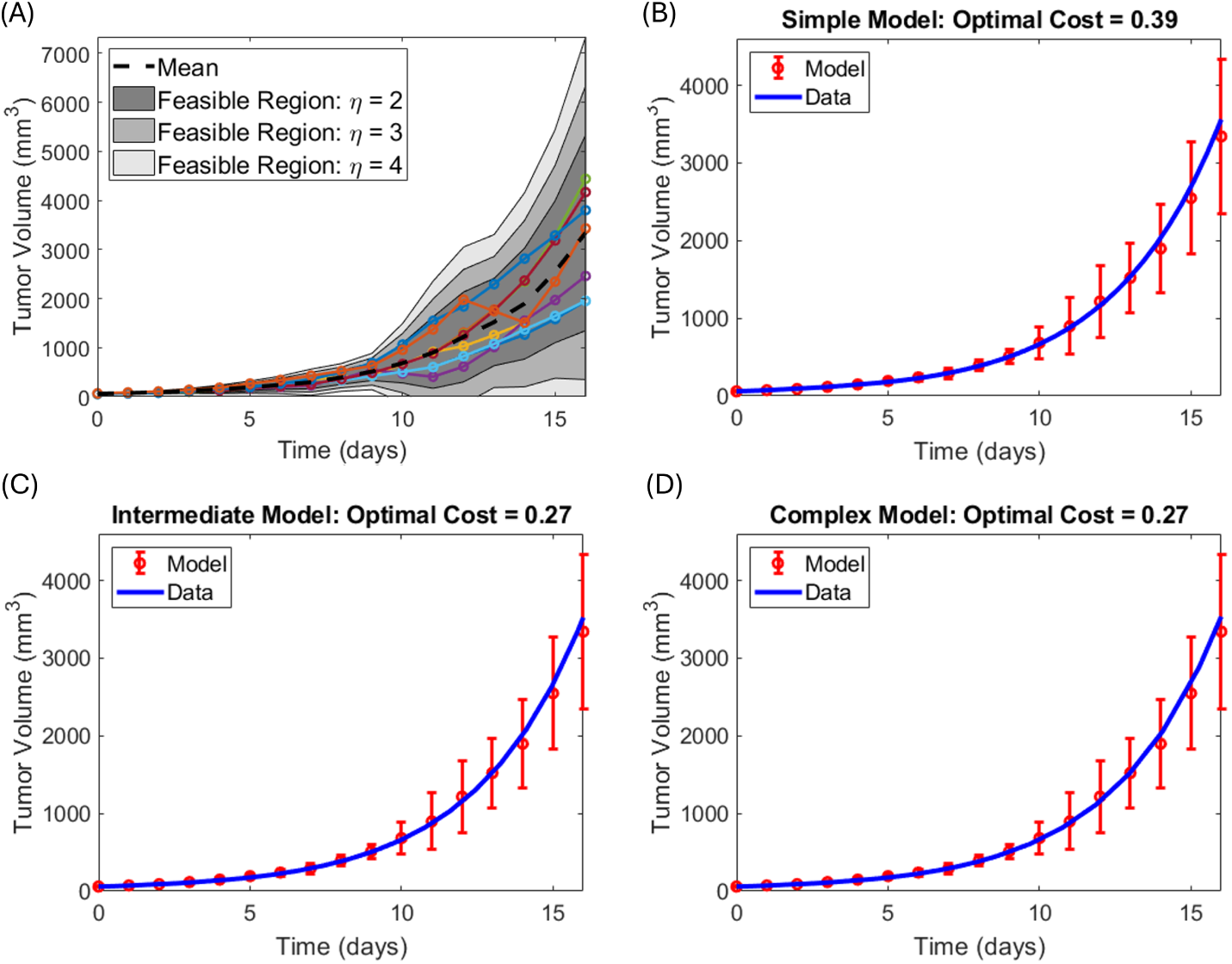
Volumetric time course of murine melanoma growth subject to OV therapy. (A) Individual mouse trajectories [20] (solid colored curves) overlaid on top of potential feasible regions, *F*. (B-D) Average volume *±* standard deviation [20] along with model best fit. Best-fit parameters are shown in Table 1

**Table 1.** Definition of prior distributions and best-fit parameters in each of the simple, intermediate and complex models. The uniform distribution *U* is defined by its lower bound *a*and upper bound *b*. The normal distribution *N* is defined by its mean *µ* and standard deviation *σ*.

| Parameter<br>(units) | $\mathcal{U}(a, b)$ | | $\mathcal{N}(\mu, \sigma)$ | | Best Fit by Model | | |
| --- | --- | --- | --- | --- | --- | --- | --- |
| | $a$ | $b$ | $\mu$ | $\sigma$ | Simple | Interm. | Complex |
| $r$ (day<sup>-1</sup>) | 0.28 | 0.37 | 0.325 | 0.015 | 0.28 | 0.28 | 0.28 |
| $\beta_m$ (vol<sup>-1</sup> day<sup>-1</sup>) | 10<sup>-3</sup> | 10<sup>-1</sup> | 0.0505 | 0.0165 | $7.63 \times 10^{-3}$ | – | – |
| $\beta_r$ (day<sup>-1</sup>) | $4 \times 10^{-4}$ | $4 \times 10^{-3}$ | $2.2 \times 10^{-3}$ | $0.6 \times 10^{-3}$ | – | $2.21 \times 10^{-3}$ | $2.27 \times 10^{-3}$ |
| $\delta_V$ (day<sup>-1</sup>) | 0.01 | 5 | 2.55 | 0.8167 | 0.595 | 4.94 | 5.00 |
| $\alpha$ | 1000 | 5000 | 3000 | 667 | – | 1900 | 1849 |
| $\gamma_I$ (day<sup>-1</sup>) | 10<sup>-5</sup> | 10<sup>-2</sup> | $5 \times 10^{-3}$ | 0.0017 | – | – | $5 \times 10^{-4}$ |
| $\epsilon$ | 0 | 1 | 0.5 | 0.1667 | – | – | 0.5025 |
| $\lambda$ (day<sup>-1</sup>) | 0 | 1 | 0.5 | 0.1667 | – | – | 0.2894 |

### 2.2 Hierarchy of Models

Herein we introduce a “simple”, “intermediate”, and “complex” model of oncolytic virotherapy. We note that this language is chosen to represent a relative hierarchy of model structures, rather than an absolute classification. The “simple” model we construct is purely phenomenological; it treats the OVs as a standard, non-replicating pharmacokinetic drug (similar to a baseline chemotherapy). The intermediate model is representative of a mechanistic viral replication model, and the complex model adds biological detail by incorporating immune-mediated tumor killing.

#### 2.2.1 Simple Model

The simplest model (Figure 2A) considers only two state variables, the tumor volume *T* (*t*) and the virion volume *V* (*t*), both measured in units mm^3^. We assume that the tumor grows at an exponential rate *r*, which is a reasonable assumption over short periods of time. We further assume that the viral infection of the tumor can be modeled using mass action kinetics with an infection rate *β_m_*, and that infection immediately clears the tumor cells. Finally, we assume that the virus is removed from the system with a decay rate of *δ_V_* . The resulting **simple model** that describes these dynamics is as follows:

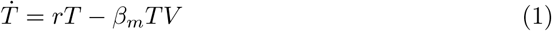

**Fig. 2.**
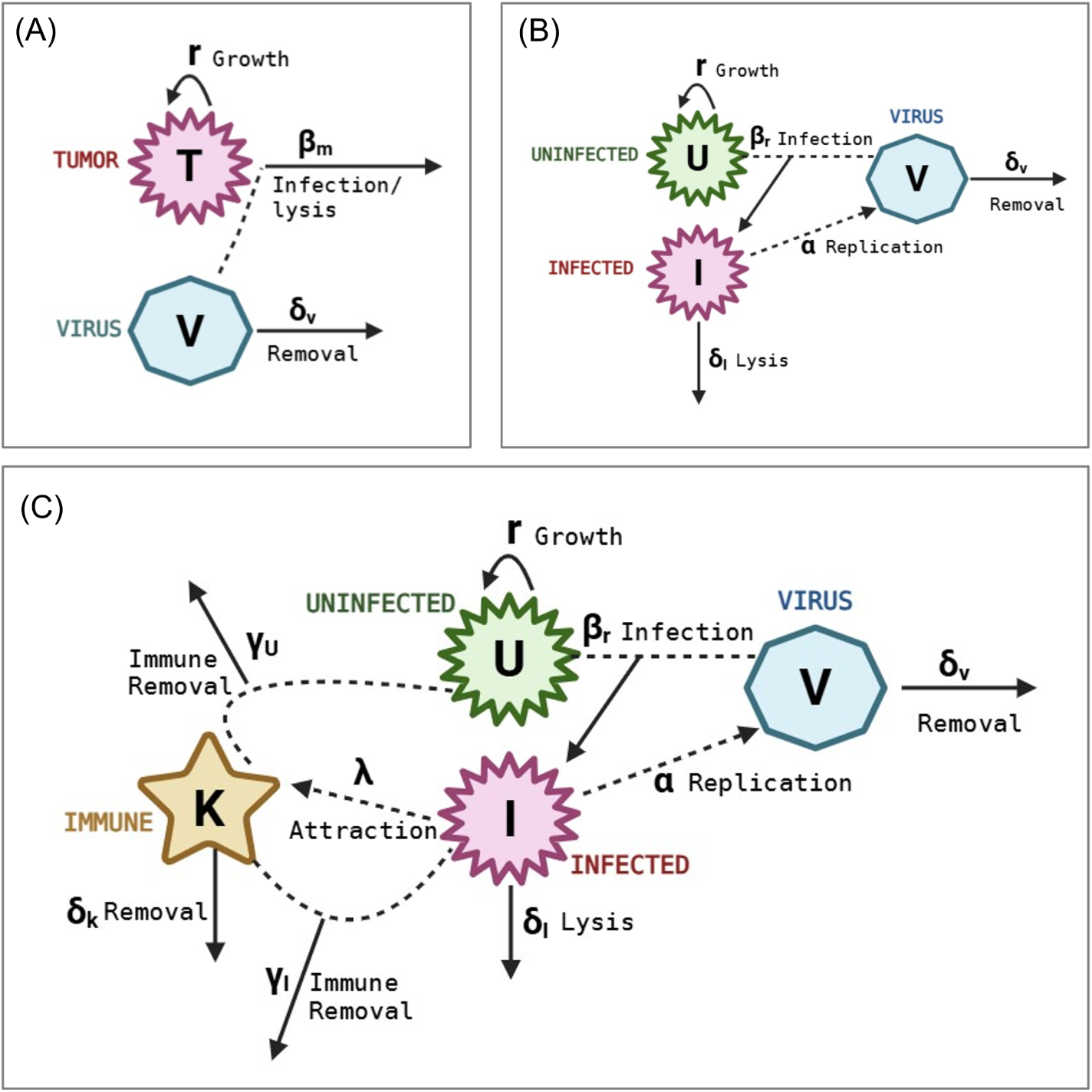
Compartment diagram for each model. (A) Simple model. (B) Intermediate model. (C) Complex model. https://BioRender.com/1oozwc7

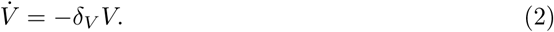

We note that this simple formulation intentionally omits known biology about oncolytic virotherapy, particularly viral burst dynamics. Instead, this simple model acts as a purely phenomenological framework that treats the therapy as a standard, non-replicating clearance drug. This simplification allows us to investigate how a model that ignores core biological mechanisms, yet still successfully fits baseline data, impacts downstream VCT outcomes.

#### 2.2.2 Intermediate Model

The intermediate model (Figure 2B) adds one variable to the simple model: the infected tumor cell population *I* (to contrast with the uninfected population *U* ). The inclusion of this one additional state variable, and two additional parameters, increases the fidelity of the model’s representation of oncolytic virotherapy. First, this allows us to more accurately model the dynamics of tumor cell death after infection. That is, infection now results in the creation of an infected cell, and it is the infected cells that are lysed and killed at the rate of *δ_I_* . In addition, the inclusion of an infected population allows for us to explicitly account for the fact that the lysis of infected cells releases *α* free virions (on average) into the tumor environment. The resulting **intermediate model** is:

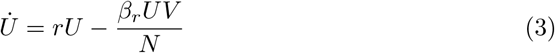

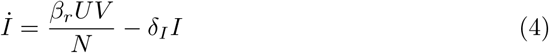

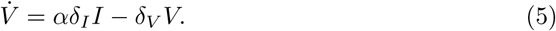

While a standard mass-action formulation offers a simpler incremental change in model complexity, we chose to implement a ratio-dependent functional response for the intermediate and complex models. This choice is motivated by the need to capture spatial constraints in solid tumors [21, 34]. By defining the total tumor cell population as *N* = *U* + *I*, the ratio-dependent transmission rate (*β_r_*) reflects that viral contact with uninfected cells is governed by their proportion within the overall tumor mass, rather than absolute cell density. (Note: *β_m_* from the simple model and *β_r_* here have different units, despite having similar interpretations.)

We also note that while the entry of a virus into an uninfected cell technically consumes a free virion, this explicit loss term is omitted here in accordance with standard modeling conventions [49, 50] and prior work [21]. Because the viral burst size is highly asymmetrical (with hundreds to thousands of virions produced per lysed cell), and because our units are volumes and not counts, the singular viral loss during infection is mathematically negligible and does not alter the system’s dynamics.

#### 2.2.3 Complex Model

The complex model (Figure 2C) adds further biological detail by explicitly modeling the immune system being attracted to the tumor site due to the presence of the oncolytic virus. We model this by adding a fourth state variable *K*(*t*), which tracks the amount of immune cells at the tumor site due to the OV. We assume that both uninfected and infected tumor cells are eliminated by these immune cells at ratio dependent rates, *γ_U_* and *γ_I_* , respectively. Given that infected cells could presumably be targeted for both the expression of tumor and viral antigens, we assume *γ_I_ ≥ γ_U_* . This can alternatively be expressed as *γ_U_* = *ɛγ_I_* , where 0 *≤ ɛ ≤* 1.

We assume the immune cells are recruited by viral-infected tumor cells at rate *λ* and naturally decay at a rate of *δ_K_*. After including these assumptions, we arrive at the **complex model**:

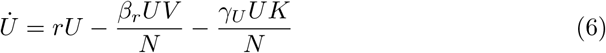

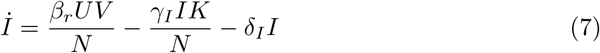

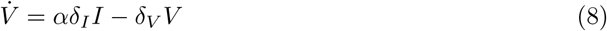

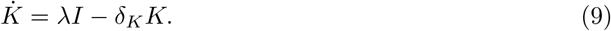

As in the intermediate model, *N* = *U* + *I*. This model has one more state variable than the intermediate model and four more parameters.

### 2.3 Model Parameter Ranges and Initial Conditions

In each model, we fix the initial condition for the (uninfected) tumor volume to be the average of the murine tumor volume at Day 0. This corresponds to using *U* (0) = 59.2 mm^3^. Any model with an infected cell compartment is initialized with zero infected cancer cells. Any model with an immune cell population is initialized with zero immune cells; note that this assumes that all immune infiltration that we are modeling is attributable to the OV therapy.

To fit the model to the data in Figure 1A, we start by establishing reasonable parameter ranges. We first focus on the parameters shared across all three models: the intrinsic growth rate *r*, the two variants of the viral infection rate *β*, and the viral decay rate *δ_V_* . Regarding the intrinsic tumor growth rate, data suggests that the tumor cell doubling time is between 2 to 3 days [51], which corresponds to *r ∈* [ln 2*/*3, ln 2*/*2] *≈* [0.23, 0.35]. Individually fitting an exponential growth model to the control mice in the trial resulted in *r ∈* [0.28, 0.37] (see Figure S1), so we chose to use this as the allowable domain for *r*.

For the transmission rate, we wanted a wider variety of values, as experimental data is difficult to ascertain. We allowed *β_r_* to range from 4 *×* 10*^−^*^4^ to 4 *×* 10*^−^*^3^ vol*^−^*^1^ day*^−^*^1^ whereas *β_m_* ranged from 10*^−^*^3^ to 10*^−^*^1^ per day. The virus removal rate *δ_V_* was restricted to be between 0.01 and 5 per day [21, 32, 34] to capture values seen throughout the literature and allow for expanded values on both sides of the range.

The intermediate model introduces a lysis rate for infected cells and a viral burst size. Given the tightly coupled relationship between these parameters, we chose to fix the infected cell lysis rate to *δ_I_* = 1 per day and then constrain *α* between 1000 and 5000 viruses [52]. Finally, the complex model introduces parameters related to immune cell infiltration and tumor-kill. We constrain *γ_I_* between 10*^−^*^5^ and 10*^−^*^2^ per day and *λ* between 0 and 1 per day. As *ɛ* is the scaling factor that determines the immune system’s impact on uninfected cells relative to infected cells, 0 *≤ ɛ ≤* 1. We fixed *δ_K_* = 0.35 per day. The parameter ranges used for all models are summarized in Table 1.

### 2.4 Plausible Population Generation

To generate plausible populations (PPops), we need to establish an underlying distribution for each model parameter (a prior distribution) and a method for including/excluding a sampled parameterization in the PPop (to determine the parameter posterior distribution). Importantly, the algorithms implemented herein serve strictly to isolate biologically feasible parameter sets (PPops); they do not generate a statistically weighted virtual population (VPop) calibrated to the demographic distributions of a specific clinical trial [53], which is outside of the scope of this work given the data available from the preclinical OV trial [20].

Herein, we consider two commonly used approaches for generating plausible populations, accept-or-reject and accept-or-perturb [18]. These methods are summarized in Figure 3. Accept-or-reject is a form of Approximate Bayesian Computation [54, 55], where a parameterization is accepted whenever its corresponding model trajectory sufficiently “matches” the data. Herein, we define the feasible region for a plausible patient as follows:

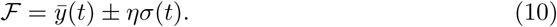

**Fig. 3.**
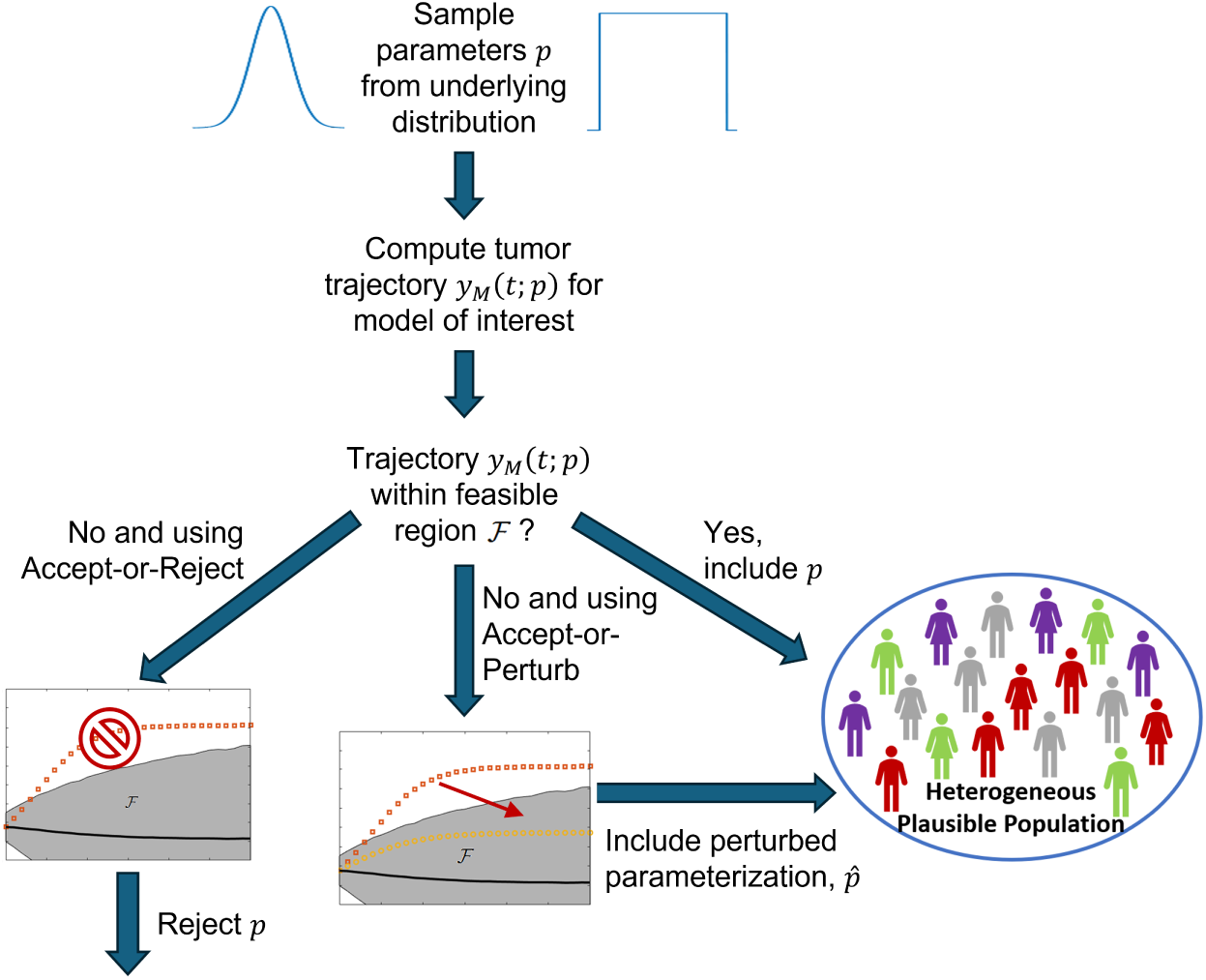
Summary of accept-or-reject and accept-or-perturb methods for generating plausible patient populations.

Thus, *F* represents a region around the mean patient trajectory *̄y*(*t*), spanning *η* standard deviations at each time point *t*. Any parametrization producing a trajectory that deviates outside *F* at any time point is discarded. This definition results in a feasible region that is quite narrow at early time points and widens as time progresses, consistent with how the tumor volume changes over time in the experimental data (Figure 1A).

We note that the plausible population is shaped by the choice of the distance threshold (*η*), as shown in Figure 1A. In prior work [18], we demonstrated that tighter thresholds (*η ≤* 2) artificially truncate the feasible region, resulting in an overly homogeneous plausible population that excludes clinically valid trajectories. Here, we see that *η* = 2 is just large enough to contain each individual mouse trajectory (Figure 1A). This choice of *η* effectively means the plausible population cannot be any more diverse than the nine mice used in the preclinical study [20]. On the other hand, looser thresholds (*η ≥* 4) over-expand the region to include outlier dynamics that deviate substantially from the data. Here, *η* = 4 broadens the feasible region so significantly that complete responders (for which the end tumor volume is below 1 mm^3^) are included in the plausible population (Figure 1A). Given that all mice in the preclinical study exhibited progressive disease (end volume at least 20% greater than initial volume), the use of *η* = 4 is overly permissive. This is consistent with our previous analysis [18] that found that *η* = 3 is a “sweet spot” that allows for the inclusion of valid trajectories without introducing non-representative behaviors. Further, assuming a normal distribution of tumor volumes, *η* = 3 theoretically captures 99.7% of the target distribution. This results in a feasible region that is wide enough to capture realistic inter-patient heterogeneity without including extreme outliers. Thus, we hold this threshold constant at *η* = 3 in the present study, unless otherwise indicated.

The accept-or-perturb method, introduced by Allen et al. [53], does not automatically reject parameterizations that correspond to out-of-bound trajectories. Instead, parameters sampled from an underlying prior are tested for trajectory matching. Parameterizations corresponding to out-of-bound trajectories are optimized via simulated annealing (implemented herein using MATLAB’s *simannealbnd* ) to ensure the resulting volume trajectory falls within *F*. The objective function minimized during optimization is [53, 56]:

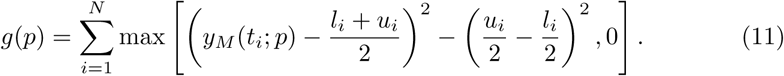

Here, *y_M_* (*t_i_*; *p*) is the model output at time *t_i_* for parameters *p*, while *l_i_* and *u_i_* denote the lower and upper bounds of the feasible tumor volume at *t_i_*. The function *g*(*p*) attains its minimum value of 0 only if all time points lie within *F*. If *g*(*p*) = 0, the parameterization is deemed a plausible patient. If *g*(*p*) *>* 0, simulated annealing perturbs *p* until the optimized parameters produce a feasible trajectory.

In the literature, it is standard to use a normal prior for accept-or-reject and a uniform prior for accept-or-perturb. In this work, we will consider both this standard pairing of prior and acceptance/rejection rule, and we will also consider what happens if we use a uniform prior for accept-or-reject and a normal prior for accept-or-perturb. Figures 4 - 7 display results when we use a normal prior for both inclusion/exclusion criteria. For a more complete comparison, see the Supplementary Information which contains comparisons for all four combinations of priors and inclusion/exclusion criteria. The bounds of the uniform prior are chosen to precisely match the parameter ranges established in Section 2.3. The mean and standard deviation of the each normal prior is computed so that the bounds of the corresponding uniform prior for that parameter are exactly three standard deviations from the mean (see Table 1). Note that we assume that the parameters that vary across plausible patients are pairwise independent. Thus, each prior is represented by a univariate distribution. Each plausible population will contain 500 plausible patients, as prior work has shown that VCT results remain relatively consistent for population sizes of 500 or larger for models of the relative complexity considered in this work [18].

**Fig. 4.**
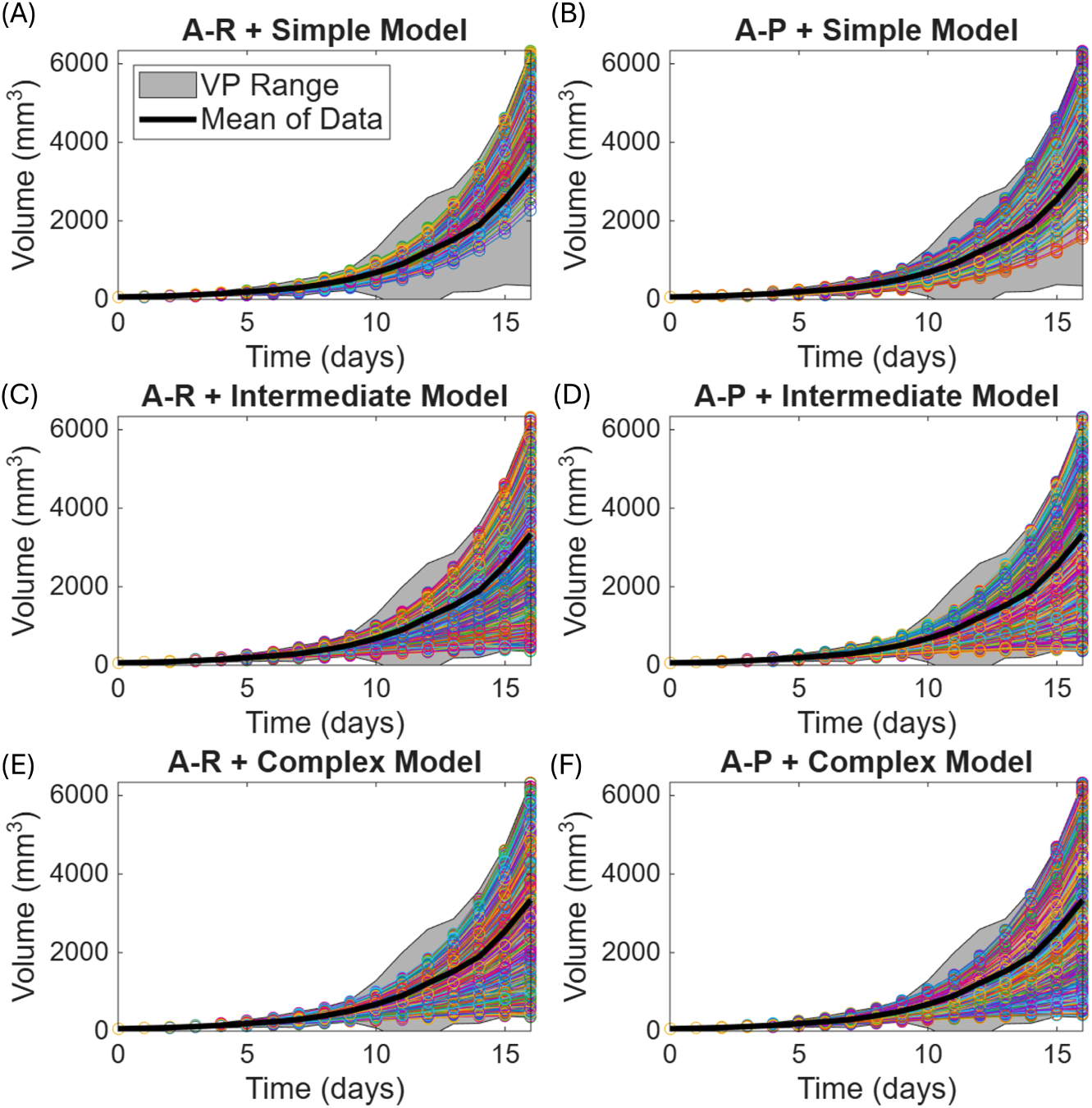
Extent to which plausible populations span the feasible trajectory space when using a normal prior. (A)-(B) Simple model. (C)-(D) Intermediate model. (E)-(F) Complex model. The first column containing (A), (C) and (E) shows the results using accept-or-reject (A-R), and the second column containing (B), (D) and (F) shows the results using accept-or-perturb (A-P). The associated violin plots and uniformity scores (*D*) for these cases and with a uniform prior are shown in Figures S2 - S4.

### 2.5 Analysis of Plausible Populations and Virtual Clinical Trial Outcomes

The outcome of a virtual clinical trial could sensitively depend on the “patients” in the trial. We will analyze how the posterior distributions (the plausible patient populations) vary across model complexity, choice of prior distribution, and choice of inclusion/exclusion criteria.

We will also conduct a virtual clinical trial on the various PPops. The trial we consider keeps the experimental protocol fixed (administering OVs on days 0, 2, 4), but varies the dose of OV given each administration day. We will measure the outcomes of each trial by computing the percent of plausible patients that fall within each of the following subgroups, according to the RECIST criteria [57]:

1. Complete responders (CR): defined here as tumor volume dropping below 1 mm^3^.
2. Partial responders (PR): defined as a non-complete responder in which the tumor volume decreased by at least 30% from its baseline.
3. Progressive disease (PD): defined as at least a 20% increase in tumor volume from its baseline.
4. Stable disease (SD): defined as any response not classified as CR, PR, or PD.

In all cases, tumor volume is assessed at the end of the experimental time window, which corresponds to 16 days post treatment initiation.

## 3 Results and Discussion

### 3.1 Model Fits to Data

We first seek to establish that each of the simple, intermediate, and complex models can well-describe the average tumor volume time-course data. Each model is fit to the OV trial data using a multi-start fitting algorithm implemented in MATLAB. First, *N* points are randomly sampled from the domain defined for the uniform priors (Table 1) using Sobol sampling. Next, the function *fmincon* is used to minimize the normalized (by the variance in the data at each time point, *σ*^2^(*t_i_*)) sum of the the square error between the model, *y_M_* (*t_i_*; *p*), and the data, *y_data_*(*t_i_*):

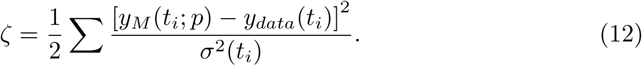

Figures 1(B)-(D) show the best fits of the simple, intermediate, and complex model, respectively, to the experimental data. The best-fit parameter values that result from minimizing *ζ* are given in Table 1. We observe that all three models can well-describe this experimental data, with the intermediate and complex model having a slightly lower optimal cost than the simple model.

### 3.2 Plausible Population Variability Across Models

To be accepted as a plausible patient, the choice of parameters defining the patient must result in a solution to the mathematical model that stays within *η* = 3 standard deviations of the mean of the trajectory defined by the experimental data, as defined in equation (10). Figure 4 (normal prior) and Figures S2 - S4 show how the accepted PPops cover this feasible region as a function of model complexity and prior/inclusion criteria pairing.

We observe that the trajectories corresponding to the simple model span the smallest fraction of feasible trajectory space, with accept-or-perturb (Figure 4B) having somewhat better coverage than accept-or-reject (Figure 4A). However, as model complexity increases from the intermediate model (Figure 4C-D) to the complex model (Figure 4E-F), little difference is observed in the coverage of feasible trajectory space. This can also be seen in the violin plots in Figure S2, which shows the distributions of tumor volumes at four time points across six settings: the three models paired with accept-or-reject and accept-or-perturb using a normal prior. As in Figure 4, tumor volumes are similarly constrained by Day 4, but diverge significantly by Day 16. Specifically, the simple models maintain narrower, more concentrated distributions, indicating a tighter constraint on the predicted trajectory space. In contrast, the intermediate and complex models exhibit much broader, top-heavy distributions by Day 16. This reinforces that increased model complexity expands the span of feasible trajectories while yielding minimal differences between the intermediate and complex models. The violin plots in Figure S2 also demonstrate that accept-or-perturb produces bimodal tumor volume distributions, while the accept-or-reject results in more unimodal distributions. Similar results are seen when using a uniform prior see Figure S3.

To quantify the dispersal of the trajectories in the feasible space, we use a distance metric between probability distributions referred to as the Earth mover’s distance (EMD) [58]. To this end, denote the endpoints of each plausible trajectory (tumor volume at the final time point) as a set of points *x*_1_*, x*_2_*, . . . , x_N_ ∈ R*, where *N* is the number of plausible patients in the PPop. We want to compare the distribution of these points to the uniform distribution (*U* ) on the same interval; this will serve as a measure of dispersal of the points. The last time point of the feasible region has a lower bound of *a* = 347.353 and upper bound of *b* = 6336.03. Thus, we will compare the empirical distribution of *{x_i_}* to *U* (*a, b*).

To simplify computations, rescale the points using *y_i_* = (*x_i_ − a*)*/*(*b − a*) *∈* [0, 1]. Let *F_Y_* be the empirical cumulative distribution of *{y_i_}* and let *F_U_* = *y* be the cumulative distribution associated with *U* (0, 1). The EMD is the 1-norm distance (the 1-Wasserman distance) between the corresponding distribution functions [58]:

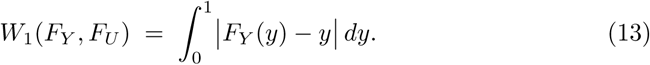

In the case that *y_i_* is uniformly distributed, *W*_1_ = 0. In the case that all points collapse at one boundary of the region, *W*_1_ = 0.5. We can thus define a scale-free quantity:

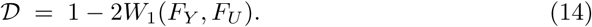

This uniformity score satisfies *D ∈* [0, 1], with *D* = 1 for uniformly spaced points (maximal dispersal), and *D ≈* 0 when all points collapse into a tight cluster (minimal dispersal).

The uniformity score *D* is computed for all three models, using both priors (uniform and normal) and inclusion/exclusion criteria (accept-or-reject and accept-or-perturb). These values, shown in Figure S4 confirm what we see visually in Figure 4 when a normal prior is used: the simple model is least dispersed throughout the feasible region, with *D* values between 0.51 and 0.54 independent of the inclusion/exclusion criterion used. Still considering the normal prior, the intermediate (*D* = 0.92 using both methods) and complex models (*D* = 0.91 using accept-or-reject and *D* = 0.88 using accept-and-perturb) result in plausible populations that comparably span the feasible region. As these uniformity scores are very close to 1, this indicates that both the intermediate and complex models result in trajectories that are near uniformly distributed in the feasible region by Day 16. These qualitative results are unchanged when using a uniform prior (see Figure S4). The impact of *η*, which determines the extent of the feasible region, on this uniformity score is explored in Figure S5.

Next we compare the resulting PPop parameter distributions for the three models under different inclusion/exclusion criteria. The results for the simple model with a normal prior are shown in Figure 5; results for both the normal and uniform prior are shown in Figure S6. We observe the posteriors generated from normal priors maintain features of a normal distribution, with the effects being more pronounced when accept-or-reject is used as the inclusion/exclusion criterion than when accept-or-perturb is used (Figure 5). Posteriors generated from a uniform prior also tend to maintain features of the uniform prior, with the notable exception of the tumor growth rate *r*, which shows a strong bias towards lower growth rates (Figure S6). This result can likely be explained by the fact that the best-fit value of the growth rate equaled the lower bound we imposed on the parameter (Table 1).

**Fig. 5.**
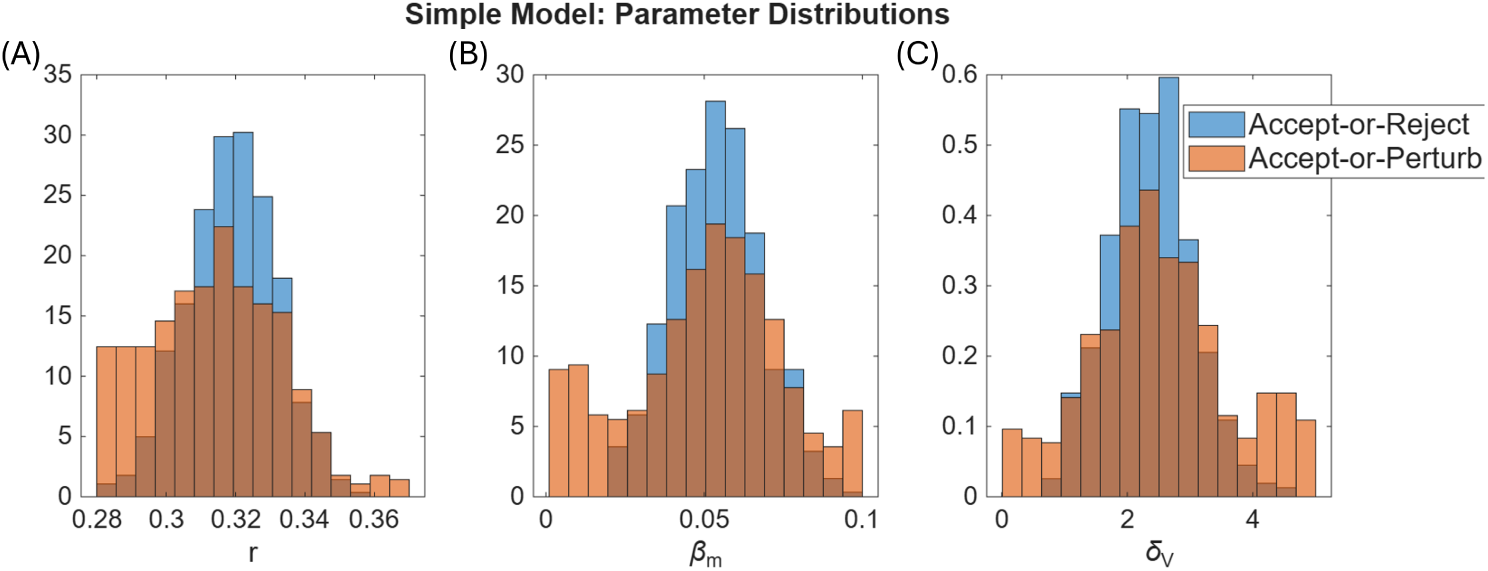
Posterior distributions for the parameters included in the plausible population using the simple model. (A) Tumor growth rate, *r*. (B) Viral infection rate, *β_m_*. (C) Viral decay rate, *δ_V_* . Results are shown for each inclusion/exclusion criterion with a normal prior distribution. Results using a uniform prior are shown in Figure S6.

The preservation of normality in the posteriors that result from using accept-or-reject is also seen with the intermediate model (Figure 6) and the complex model (Figure 7), though there are more noticeable shifts to a left-or right-tailed distribution in these more detailed models than in the simple model. For accept-or-perturb, using both the intermediate and complex models, the posterior distributions do not mimic the normality seen in the prior distributions (Figures 6 and 7). When the uniform prior is used with both accept-or-perturb and accept-or-reject, we see that many of the parameterizations accumulate at the boundaries imposed on parameter space (Figures S7 and S8), an effect that has been reported previously [19, 59].

**Fig. 6.**
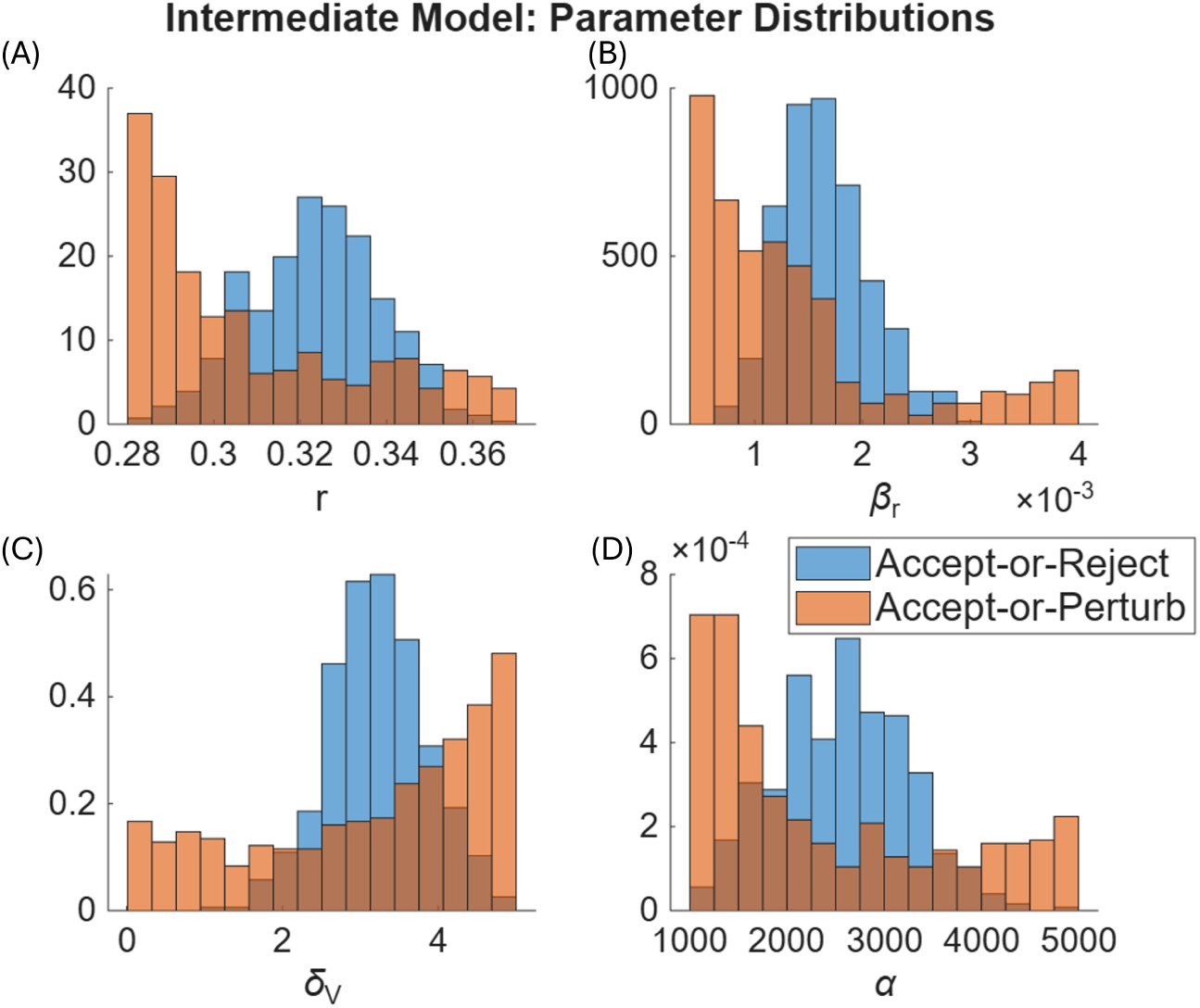
Posterior distributions for the parameters included in the plausible population using the intermediate model. (A) Tumor growth rate, *r*. (B) Viral infection rate, *β_r_*. (C) Viral decay rate, *δ_V_* . (D) Viral burst size, *α*. Results are shown for each inclusion/exclusion criterion with a normal prior distribution. Results using a uniform prior are shown in Figure S7.

**Fig. 7.**
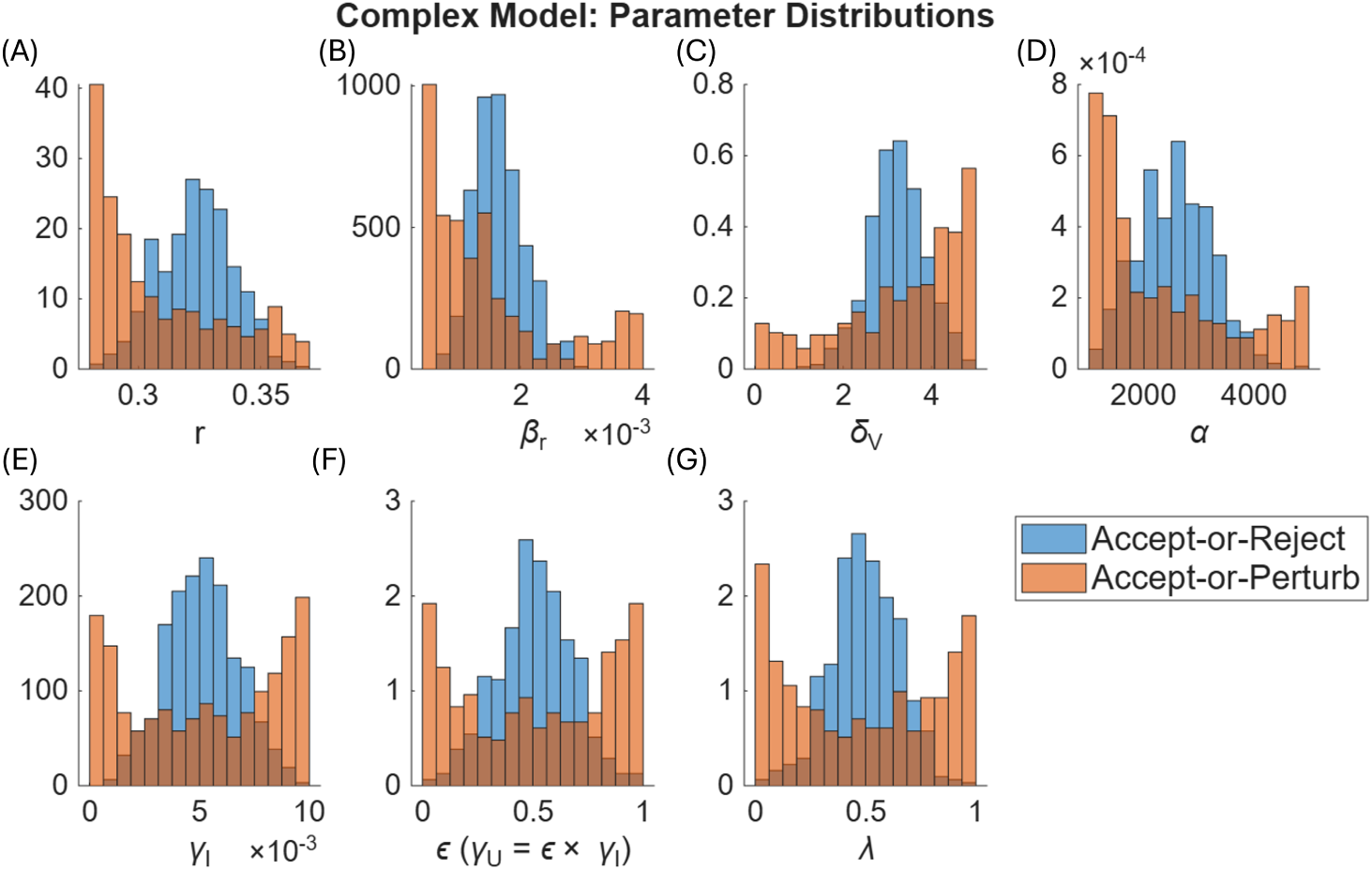
Posterior distributions for the parameters included in the plausible population when the population using the complex model. (A) Tumor growth rate, *r*. (B) Viral infection rate, *β_r_* . (C) Viral decay rate, *δ_V_* . (D) Viral burst size, *α*. (E) Immune-kill rate of infected cell, *δ_I_* . (F) Scaling factor for immune-kill rate of uninfected cells, *ɛ* where *δ_U_* = *ɛδ_I_* . (G) Immune cell recruitment rate, *λ*. Results are shown for each inclusion/exclusion criterion with a normal prior distribution. Results using a uniform prior are shown in Figure S8.

To further explore the preservation of prior distribution characteristics on posterior distributions, we generated posterior distributions using uniform prior distributions, as is standard for accept-or-perturb (Figures S6 - S8). To quantify the difference in posterior distributions, we calculated the sum of the absolute difference in the normalized histogram values for each parameter. Dividing this quantity by 2 results in a normalized metric in which a value of 0 indicates identical histograms and a value of 1 indicates maximally distinct histograms.

Visually, the accept-or-perturb method results in posterior distributions that appear less sensitive to whether a uniform or normal prior was used (Figures S6 - S8). The distance metric confirms these observations. In particular, for the simple model, the distance metric for accept-or-perturb ranged from 0.224 to 0.286. When accept-or-reject is used, the difference measure ranged from 0.372 to 0.406, indicating larger discrepancies between the posterior distributions based on the choice of prior.

Values of the distance metric using accept-or-perturb ranged from 0.066 to 0.126 for the intermediate model and 0.088 to 0.136 for the complex model. As these values are smaller than for the simple model, this demonstrates that PPops generated from models of sufficient complexity are less influenced by the prior than simple models when using accept-or-perturb.

The same cannot be said, however, when using accept-or-reject, as we observe comparable distance metrics independent of model complexity. In particular, the metric ranges from 0.368 to 0.446 for the intermediate model and 0.370 to 0.446 for the complex model. Overall, we conclude that the prior distribution had a much larger effect on the resulting posterior distribution when using the accept-or-reject as compared to accept-or-perturb. Using accept-or-perturb with models of “sufficient” detail mitigates the bias imposed by the prior.

### 3.3 Comparison of Trial Results

At the experimental dose, the individual mice exhibited no qualitative difference in treatment response (Figure 1A) - all mice demonstrate progressive disease. Thus, we designed a virtual clinical trial to study the impact of increasing the viral dose, from a minimum value of 10 *×* 10^9^ (the dose used in the experiments) to a maximum value 400 *×* 10^9^ virions per dose. Otherwise, the protocol design is unchanged, meaning drug is still administered on the same days as in experiments (Days 0, 2, 4).

The resulting trial outcomes and the dependence on trial design (model type, prior, and inclusion/exclusion criteria) are shown in Figure 8. Here we compare the standard pairings of accept-or-reject with normal priors and accept-or-perturb with uniform priors, though additional pairings are shown in Figure S9A. While all trials predict the expected dose-response relationship, with higher doses yielding more complete responders, quantitative differences are observed across dosing space depending on trial design. Focusing on the CR group (Figure 8A), trials conducted using accept-or-reject (with the normal prior) predict a homogeneous population-level response at the highest dose tested, with all plausible patients are classified as complete responders. Compare this to the PPops generated using accept-or-perturb, where only 81.6% to 92.2% of the PPop is classified as CRs. Interestingly, PPop diversity at the highest dose considered using accept-or-perturb actually decreases as a function of model complexity, with the simple model exhibiting a more diverse response than the complex model.

**Fig. 8.**
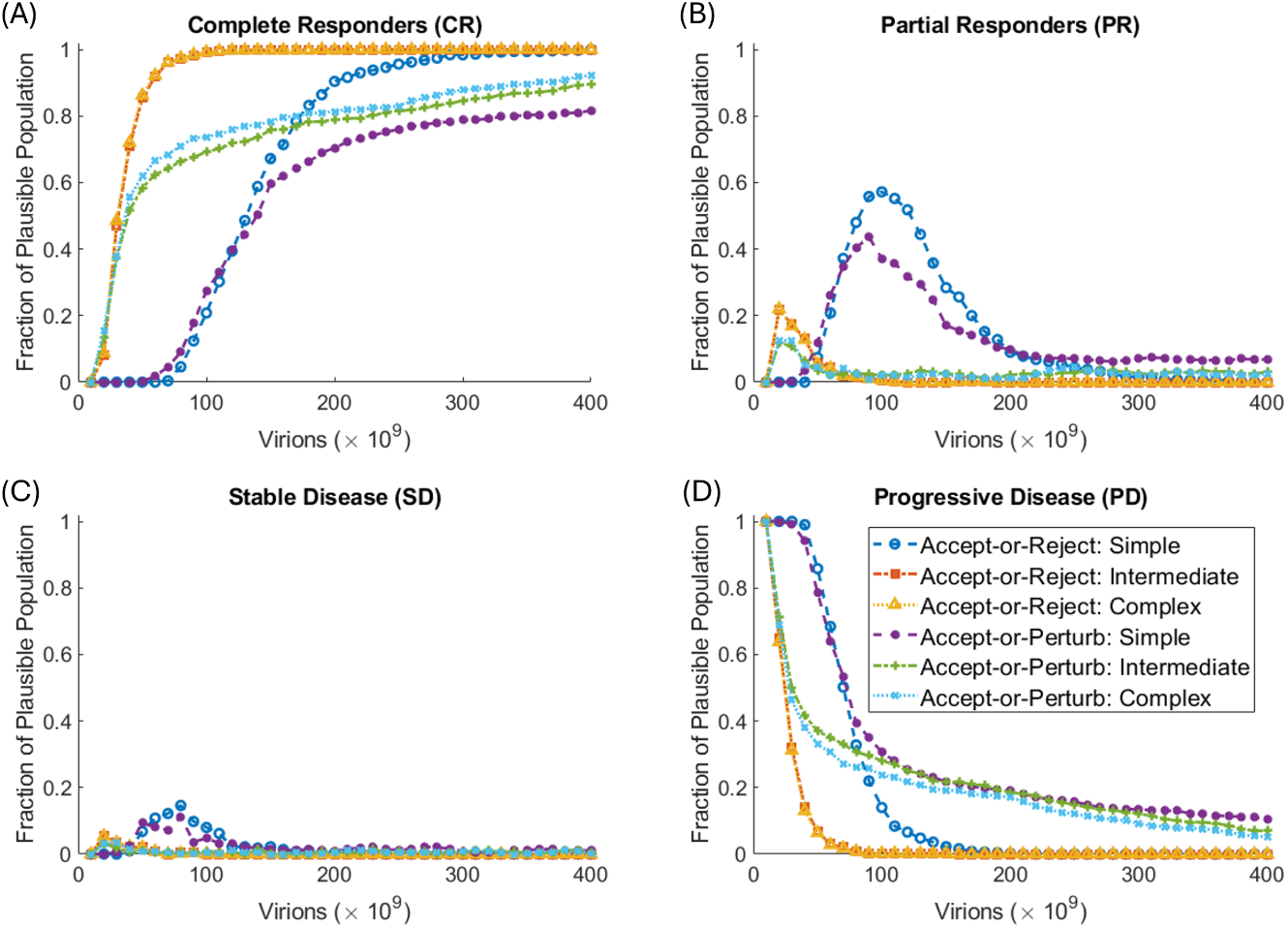
Virtual clinical trial outcomes for the simple, intermediate and complex models using accept-or-reject with a normal prior and accept-or-perturb with a uniform prior. Results are shown across dosing space as the fraction of the PPop classified as (A) complete responders, (B) partial responders, (C) stable disease, (D) progressive disease.

To further explore the heterogeneity of treatment response across the PPop, we use the normalized Shannon entropy, defined as

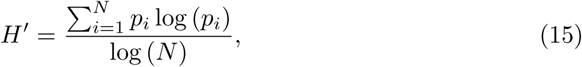

where *N* = 4 here represents the number of response categories and *p_i_* gives the probability of falling in category *i*. Note the formula is only applied for *p_i_ >* 0. Division by log (*N* ) normalizes the Shannon entropy so that minimum entropy (when all plausible patients are in one response category) corresponds to a value of 0 and maximum entropy (perfectly balanced categories) corresponds to a value of 1.

The normalized Shannon entropy across all doses considered in the trial is shown in Figure 9 for the standard pairing of prior and inclusion/exclusion criteria. See Figure S9A for the results using the “flipped” priors, and Figure S10 for how the extent of the feasible region, *η*, influences the normalized Shannon entropy. At sufficiently high doses, we still observe that the heterogeneity in trial outcomes is more sensitive to the inclusion/exclusion criteria than the complexity of the model. However, at low doses (below 50 *×*10^9^ virions), we observe the opposite: response heterogeneity is more sensitive to model complexity. In particular, heterogeneity is quite low when using the simple model and very high when using the intermediate or complex model.

**Fig. 9.**
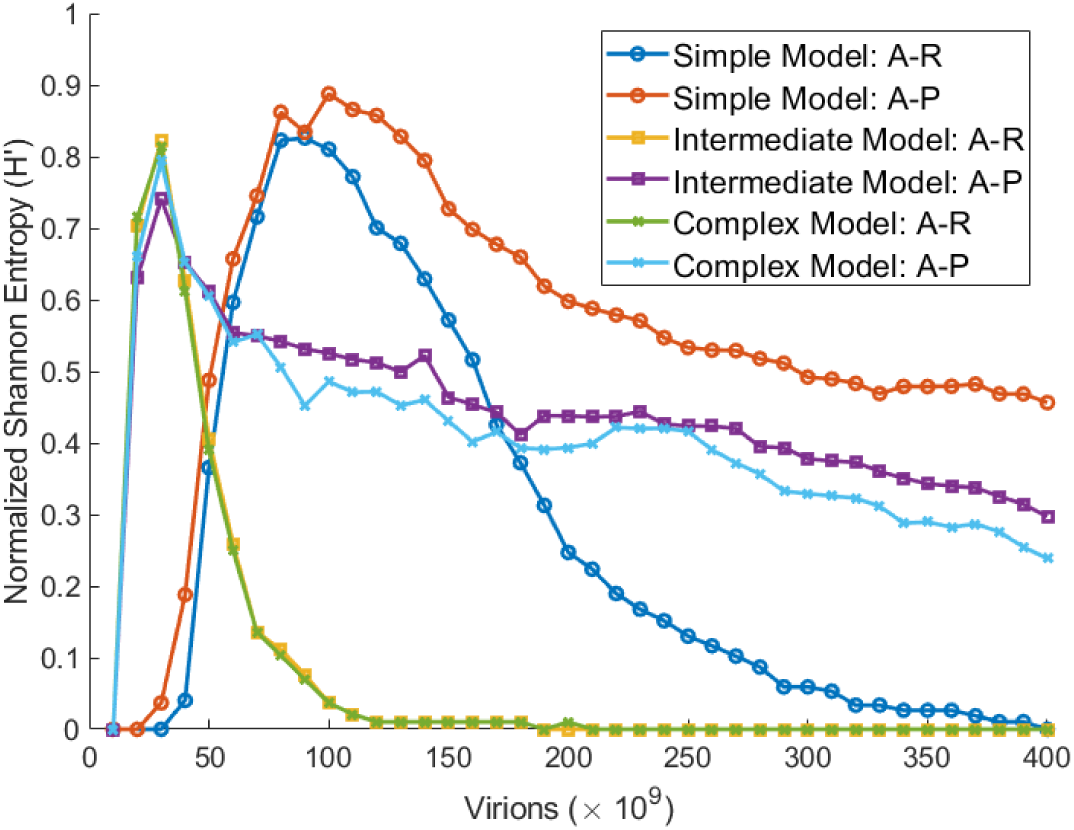
Normalized Shannon entropy of the virtual clinical trial outcomes for the simple, intermediate and complex models using accept-or-reject with a normal prior and accept-or-perturb with a uniform prior. Results for the “flipped” priors are shown in Figure S9(A).

### 3.4 Feature Selection Across Subgroups

After conducting a virtual clinical trial, there are a number of questions that can be asked about the population. For instance, one may seek to identify patient features (model parameters) that are most predictive of treatment response. Similarly, one may seek to uncover features of rare subpopulations, such as those who respond exceptionally well or poorly to treatment. It is interesting to ask how model complexity impacts the identification of these features.

Here, we will explore that question by comparing the simple model (Figure 10) with the intermediate model (Figure 11). We choose to use the intermediate model as it gives predictions very similar to the complex model. We will focus on binary response, dividing plausible patients into either those with progressive disease or those without PD (CR, PR, SD). We will consider the dose of 50 *×* 10^9^ virions, as this dose maximizes the lower bound of the normalized Shannon entropy value across trial design. To detail, *H^′^* values are between 0.36 and 0.62 at this dose, and the lower bound of *H^′^* is smaller at all other doses (see Figure 9).

**Fig. 10.**
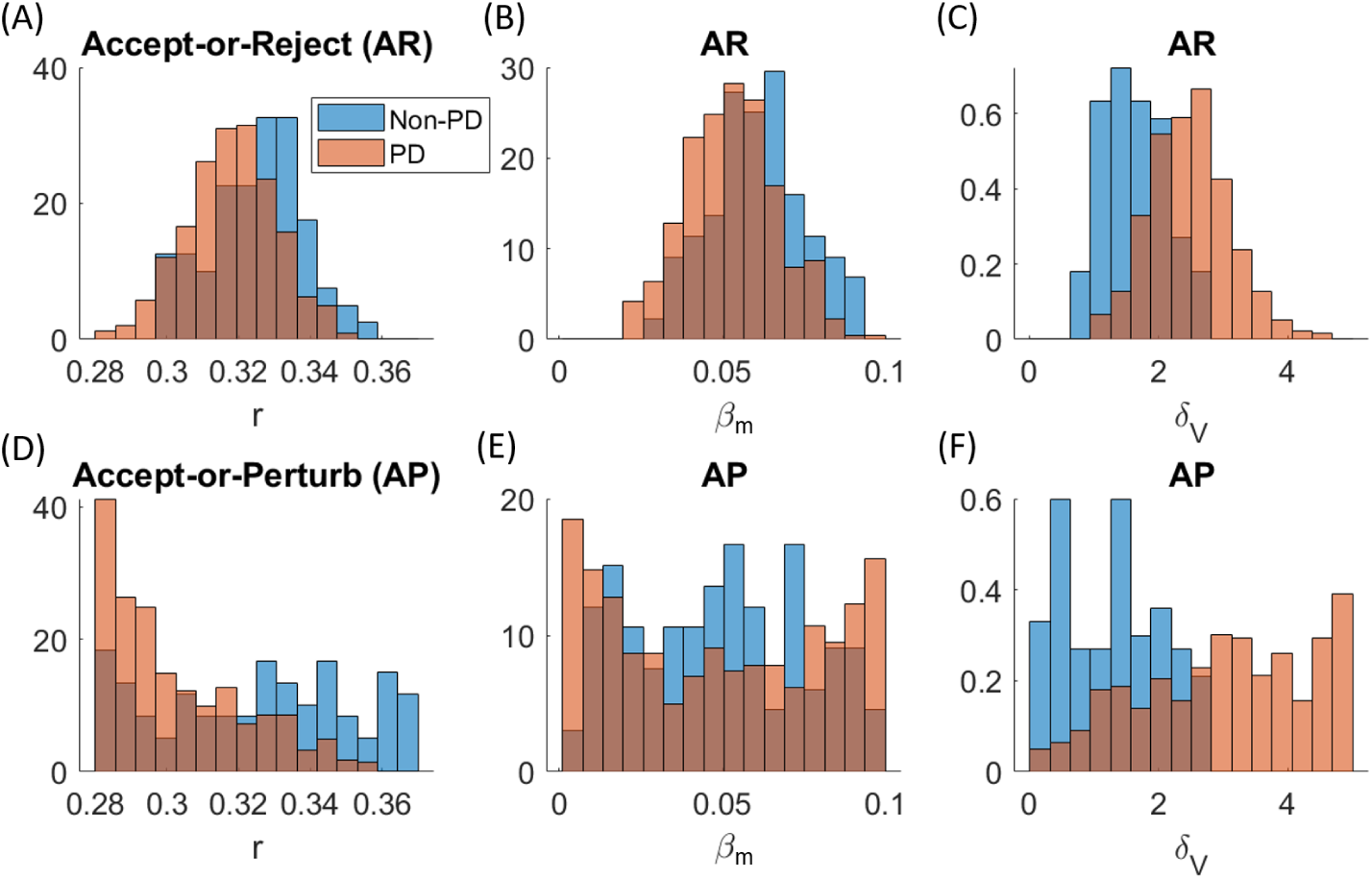
Posterior distributions in PPops generated using the simple model. (A)-(C) Posteriors using accept-or-reject with a normal prior. (D)-(F) Posteriors using accept-or-perturb with a uniform prior. PD represents the subgroup with progressive disease, and non-PD denotes all other subgroups (CR, PR, SD).

**Fig. 11.**
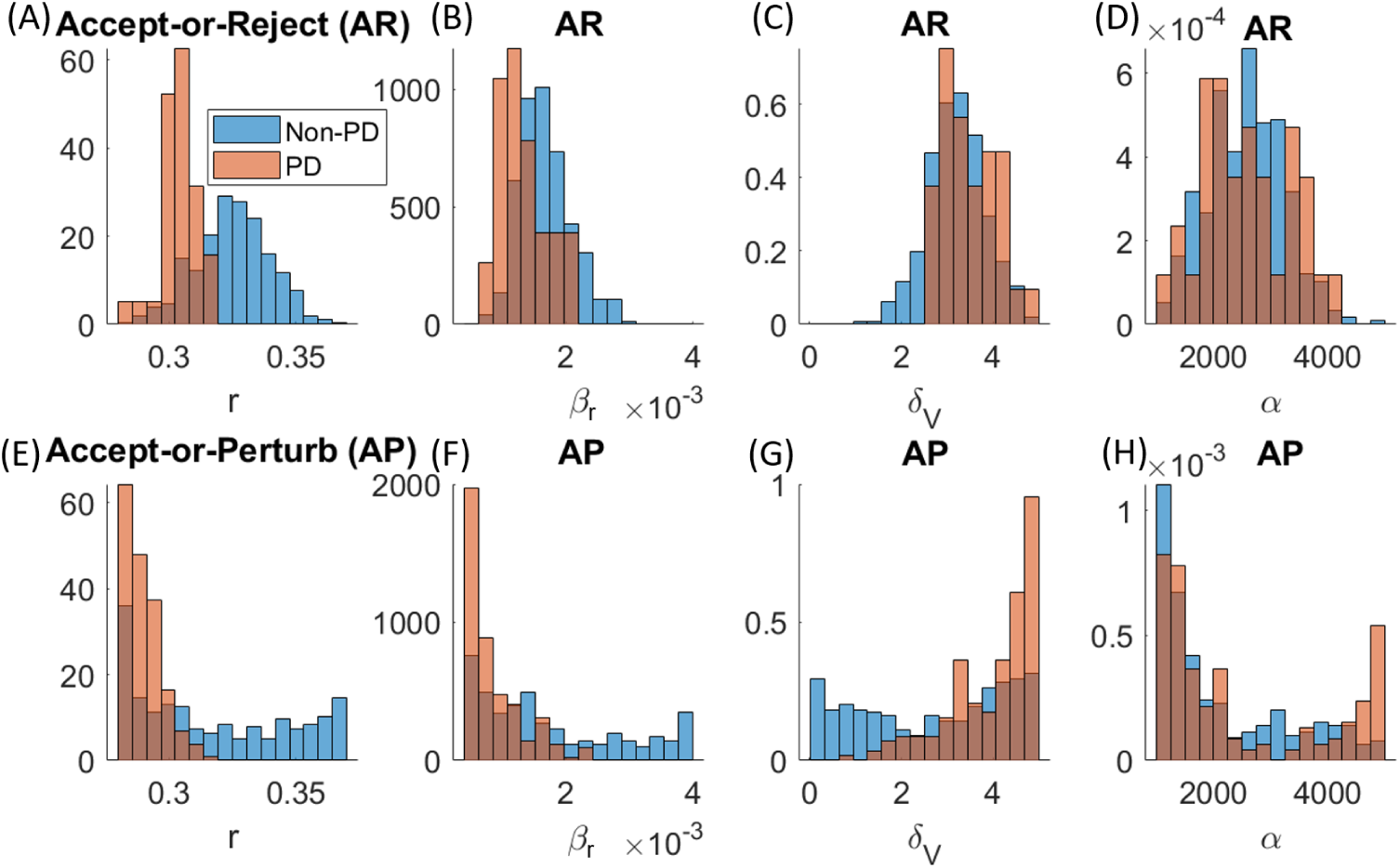
Posterior distributions in PPops generated using the intermediate model. (A)-(D) Posteriors using accept-or-reject with a normal prior. (E)-(H) Posteriors using accept-or-perturb with a uniform prior. PD represents the subgroup with progressive disease, and non-PD denotes all other subgroups (CR, PR, SD).

If we focus on the shared model parameters (tumor growth rate *r*, viral infection rate *β* and viral decay rate *δ_V_* ), we observe that progressive disease is associated with the same patient features independent of the model considered. Those with lower growth rates, lower infection rates and higher viral decay rates are more likely to be classified as non-responders. The same is true whether the PPops were generated using accept-or-reject with a normal prior (Figure 10A-C, Figure 11A-C) or accept-or-perturb with a uniform prior (Figure 10D-F, Figure 11E-G). That said, there is clearly overlap in the posterior distributions for those with PD and non-PD. A more sophisticated feature selection algorithm could help further distinguish these subgroups.

The fact that the subgroups are associated with similar features is promising, as it shows that the complexity of the model and the method for including/exclusion parameterizations may not confound feature selection associated with a specific treatment response.

## 4 Conclusions

For virtual clinical trials to live up to the promise of making clinical trials more effective and efficient, it is essential that we understand the impacts that the design of these in silico trials have on the predicted outcomes. Many decisions get made in the process of conducting a virtual clinical trial, from the selection of a mathematical model, to the determination of which parameters vary across plausible patients, to the assumed underlying distribution of these parameters, to the method that determines which parameterizations get included in the plausible population. In this work, we sought to study the role that model complexity has in virtual trial outcomes, and how that may or may not depend on parameter priors and the inclusion/exclusion criteria.

As a case study, we focused on a murine dataset of tumor progression subject to treatment with three doses of an oncolytic virotherapy. We chose this as our model system, as oncolytic virotherapy has been the subject of many modeling studies [3, 21, 26–38], with some models being biologically and mathematically simple and others being biologically detailed and mathematically complex. Our analysis shows that a model that is too simple can result in significant biases in the plausible population. This is shown in Figures 4 and S2 - S4, as we see that the plausible patient trajectories do not sufficiently span the feasible trajectory space when the simple model is used. Insufficient coverage of this allowable space likely means that the trial is analyzing a biased subgroup of the full population.

Unlike the simple model, both the intermediate and the complex models well-span the feasible trajectory space (Figures 4 and S2 - S4), meaning that trials using these models likely capture more inter-patient heterogeneity. Interestingly, little is gained in terms of coverage of trajectory space when increasing relative model complexity from the intermediate to the complex model. This suggests that once there is a sufficient level of model complexity, there may be nothing to gain from adding further complexity. This is further supported by studying the outcomes of the VCT (Figures 8, 9, S9). When we use the same prior and accept-or-reject as the inclusion/exclusion criteria, the heterogeneity in plausible patient response across dosing space is effectively the same whether we use the intermediate or the complex model. Using accept-or-perturb yields qualitatively similar results between the two models as well, though the agreement is somewhat weaker than under accept-or-reject.

That being said, there are noticeable differences in posterior parameter distributions for the plausible populations (Figures 5 - 7, S6 - S8) and in trial outcomes based on the choice of parameter priors and the inclusion/exclusion criteria (Figures 8, 9, S9). Accept-or-reject tends to result in posterior distributions that are biased by the form of the prior distribution, whereas posterior distributions that result from accept-or-perturb are less sensitive to the form of the prior. This strongly suggests that accept-or-reject, which is a standard approximate Bayesian computation approach, is insufficient for designing plausible populations. This approach too strongly biases the plausible population to resemble the prior, which may not represent clinical reality. The results of these biases are evident in the outcomes of the VCT, as there is no diversity in the response across plausible patients generated via accept-or-reject using a normal prior at sufficiently high OV doses (Figures 8 and 9). If a uniform prior is instead used (Figure S9A), there is more inter-patient variability than when a normal prior is used, but much less variability compared to accept-or-perturb. Taken together, this suggests that a sufficiently detailed model, but not too detailed, combined with accept-or-perturb is the most robust design for a virtual clinical trial.

The outcomes of this study are not without its limitations. Importantly, what we have analyzed here are plausible populations, not virtual populations. A plausible patient is a parameterization of the model that is deemed biologically feasible based on defined parameter and model output constraints. An ensemble of these biologically reasonable plausible patients forms a plausible population. If data is available to further determine how likely a plausible patient is to be in the true population, that data is utilized to identify a subset of plausible patients (virtual patients) to form a virtual population [7]. In the broader quantitative systems pharmacology (QSP) landscape, techniques have been developed to optimize this transition from plausible to virtual populations. For example, the simulated annealing framework established by Allen et al. [39] was optimized by Rieger et al. [7] through the introduction of advanced multi-stage selection algorithms designed to improve the efficiency of generating virtual populations that match clinical target distributions. When the data is available to create a virtual population (for instance [60]), some of the discrepancies that emerge in this study may be mitigated. However, when such data is not available, the impact of these design choices can be substantial.

Even if we focus only on the generation of plausible populations, another limitation of this study is that we did not consider all possible algorithms for generating these PPops, as the algorithmic landscape for VCTs is evolving quite rapidly [19, 59]. The choice of studying accept-or-reject and accept-or-perturb provides a baseline for evaluating the impact of VCT design choices and model complexity. If our VCT involved more complicated mathematical models, it would be important to consider advanced sampling architectures to overcome the computational bottlenecks and mapping limitations inherent to traditional single-chain or standard rejection frameworks. These include multi-chain adaptive frameworks like the DREAM(ZS) algorithm, which has been successfully leveraged to overcome boundary accumulation and restore hidden parameter correlation structures for high-dimensional models [19], as well as advanced sequential Bayesian extensions, including Approximate Bayesian Computation Sequential Monte Carlo (ABC-SMC) [61]. While advanced heuristic methods are important for highly complex systems, the sampling frameworks utilized in this work are sufficient to robustly map the trajectory space of our relatively low-dimensional models without adding unnecessary computational overhead.

Finally, it is important to contextualize the scale of model complexity evaluated in this study. In a translational setting, a purely phenomenological framework (like our “simple” model) is fundamentally limited in its ability to extrapolate mechanistic insights from a VCT. However, we intentionally included such a phenomenological model to study what occurs when model complexity is insufficient. There are indeed practical reasons that a model that is “too simple” could be used in a VCT. First, the principle of parsimony favors the development of the simplest model that describes the data, which ensures its parameters are structurally, and often practically, identifiable. Second, in emerging therapeutic areas in particular, a model that is “too simple” may be inadvertently utilized because the underlying physiology is not fully understood.

An analogous consideration applies to the “complex” model used in this study. While comprehensive QSP models frequently incorporate dozens of state variables and hundreds of parameters to account for intricate disease biology, our “complex” model is relatively low-dimensional by comparison. Our use of the labels “simple”, “intermediate”, and “complex” is intended as a relative hierarchy that captures the transition from a purely phenomenological clearance drug framework, to a mechanistic viral replication model, to a system incorporating immune-mediated tumor killing. Utilizing low-to-moderate dimensional models was a deliberate methodological choice. Introducing a high-dimensional QSP model would introduce even more parameter non-identifiability, which would impede our ability to determine whether changes in VCT outcomes were caused by VCT design choices and model complexity, or simply by the algorithmic challenges of sampling a high-dimensional parameter space. Furthermore, our results already demonstrate that even our “complex” model exhibits signs of over-complexity for the data.

Taken together, this work suggests that a model of intermediate complexity (enough detail to capture the biology, but not too much), coupled with accept-or-perturb as an inclusion criterion, offered the most robust VCT design approach among those we evaluated. The addition of too many biological details in the complex model did not change any of our key metrics: coverage of feasible trajectory space and heterogeneity of plausible patient response across protocol space. Furthermore, the use of accept-or-reject introduced notable biases in the plausible patient parameter posterior distributions based on the choice of prior.

While these quantitative insights are grounded in a single preclinical oncolytic virotherapy system, the fundamental structural trade-offs we observe likely extend to other contexts. That said, our findings should not be interpreted as a definitive or universal endorsement of an “intermediate” model scale, as even the definition of what makes an “intermediate” model is system-specific and subjective. Instead, this work highlights a common challenge in mathematical biology: models can easily be expanded to include endless mechanistic detail, but the data available to validate them is almost always limited. Ultimately, how to best balance model complexity against data constraints remains an open question that warrants further investigation across diverse therapeutic areas.

## Declarations

The authors have no relevant financial or non-financial interests to disclose. The code used to generate all the data presented in this manuscript is available at https://github.com/jgevertz/VCT.

## Data Availability

All data produced are available online at https://github.com/jgevertz/VCT.

## Supplementary Information

**Fig. S1.**
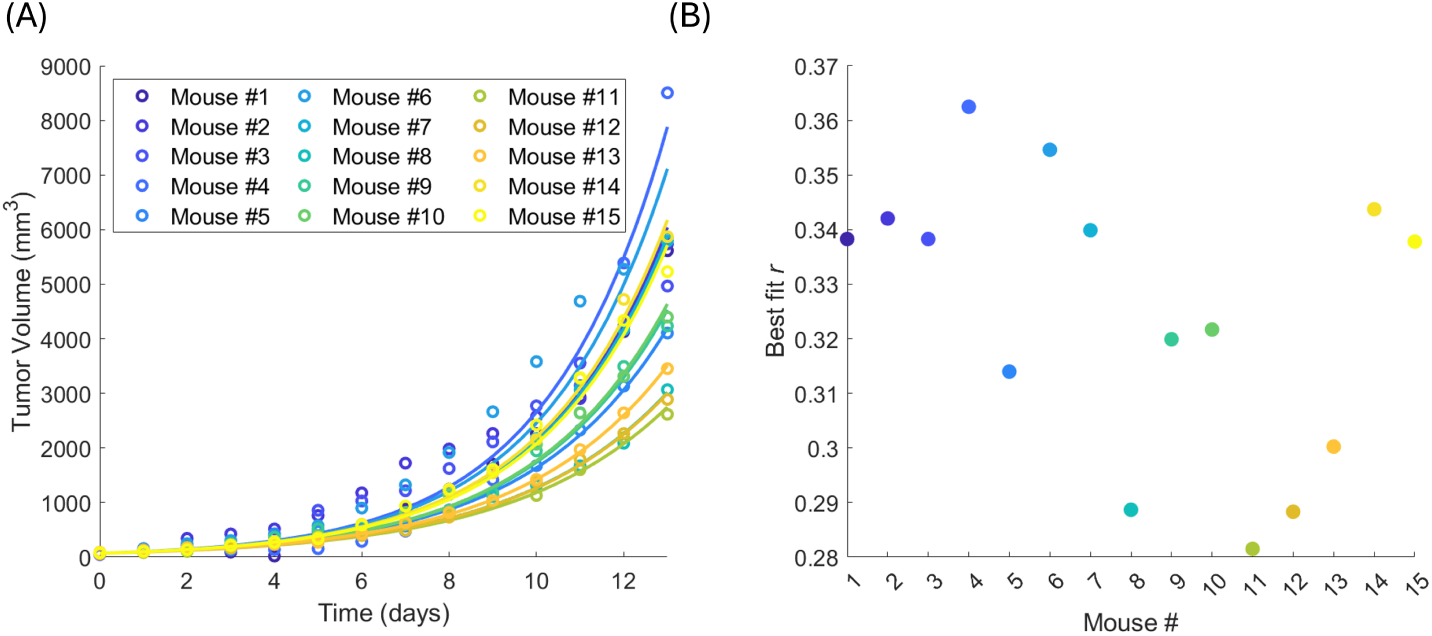
(A) Tumor volume time course for control mice (no drug) [20], along with the best fit of the equation *x*(*t*) = *x*_0_*e^rt^* to the data with *x*_0_ fixed at the average value in the control data (*x*_0_ *≈* 70.8). (B) Best-fit values of *r* for each control mouse.

**Fig. S2.**
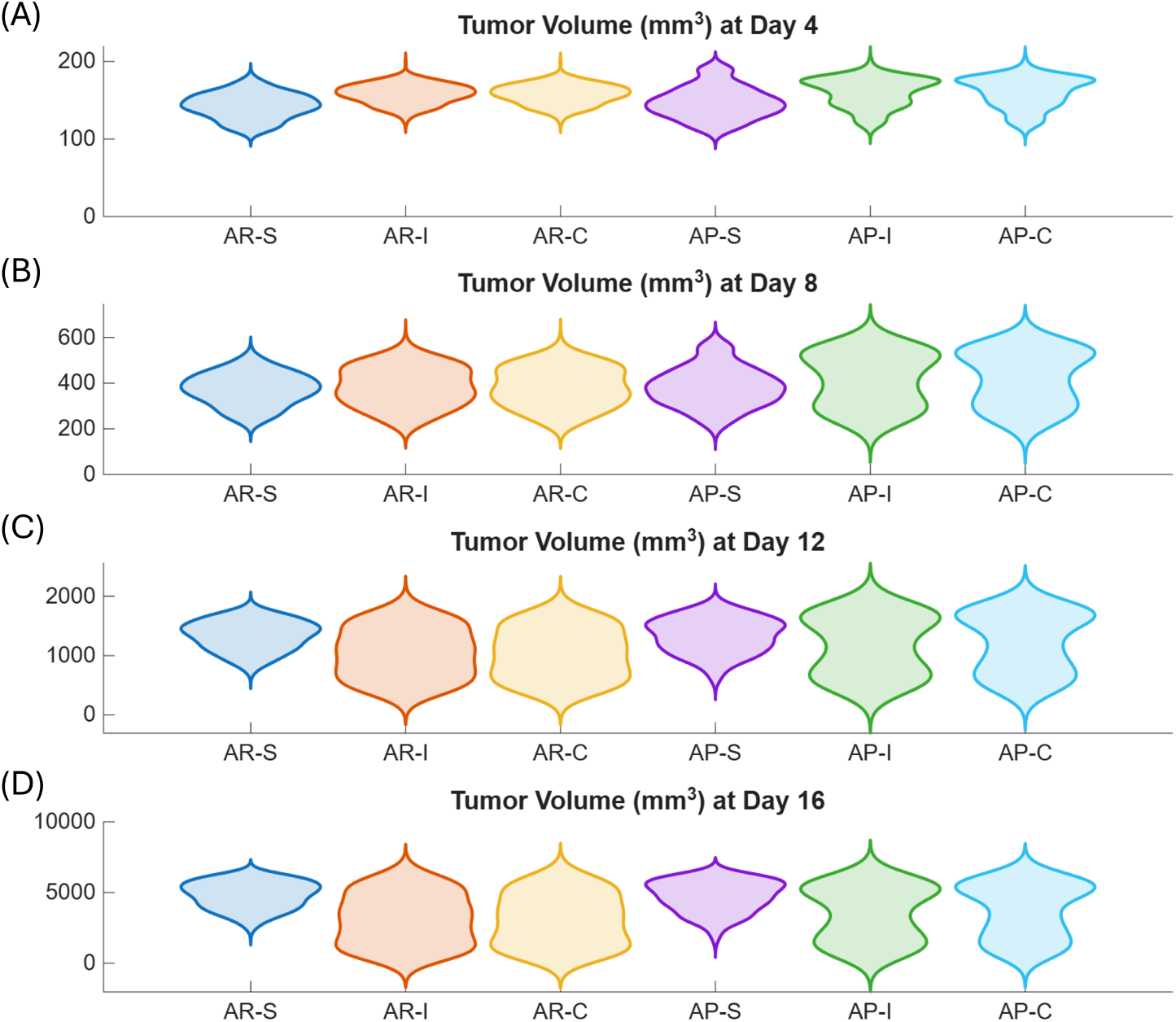
Violin plots showing the distribution and range of the plausible population within the feasible trajectory space at Days 4, 8, 12 and 16. Here, AR = accept-or-reject, AP = accept-or-perturb, S = simple, I = intermediate, C = complex. Normal prior was used.

**Fig. S3.**
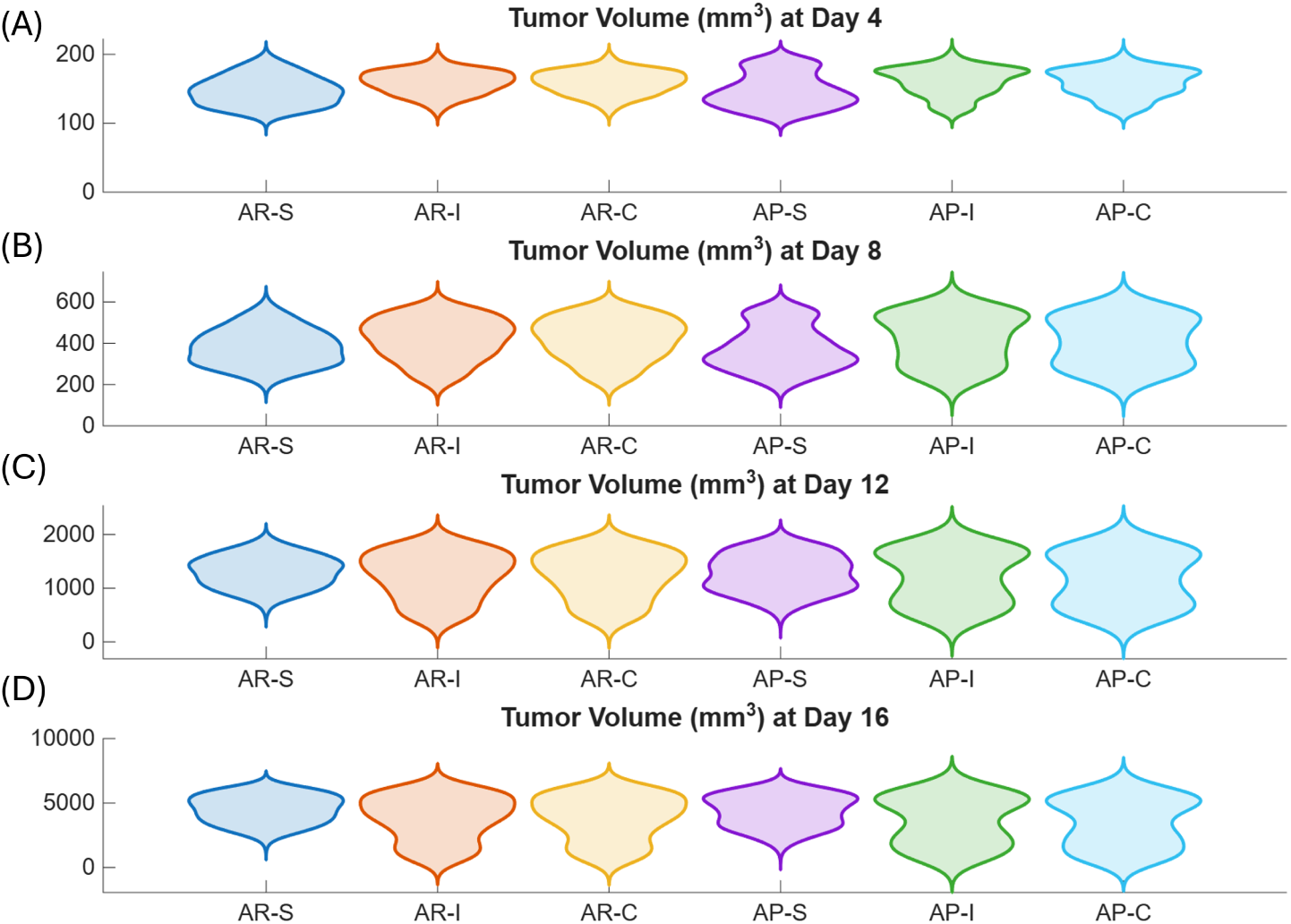
Violin plots showing the distribution and range of the plausible population within at Days 4, 8, 12 and 16. Here, AR = accept-or-reject, AP = accept-or-perturb, S = simple, I = intermediate, C = complex. Uniform prior was used.

**Fig. S4.**
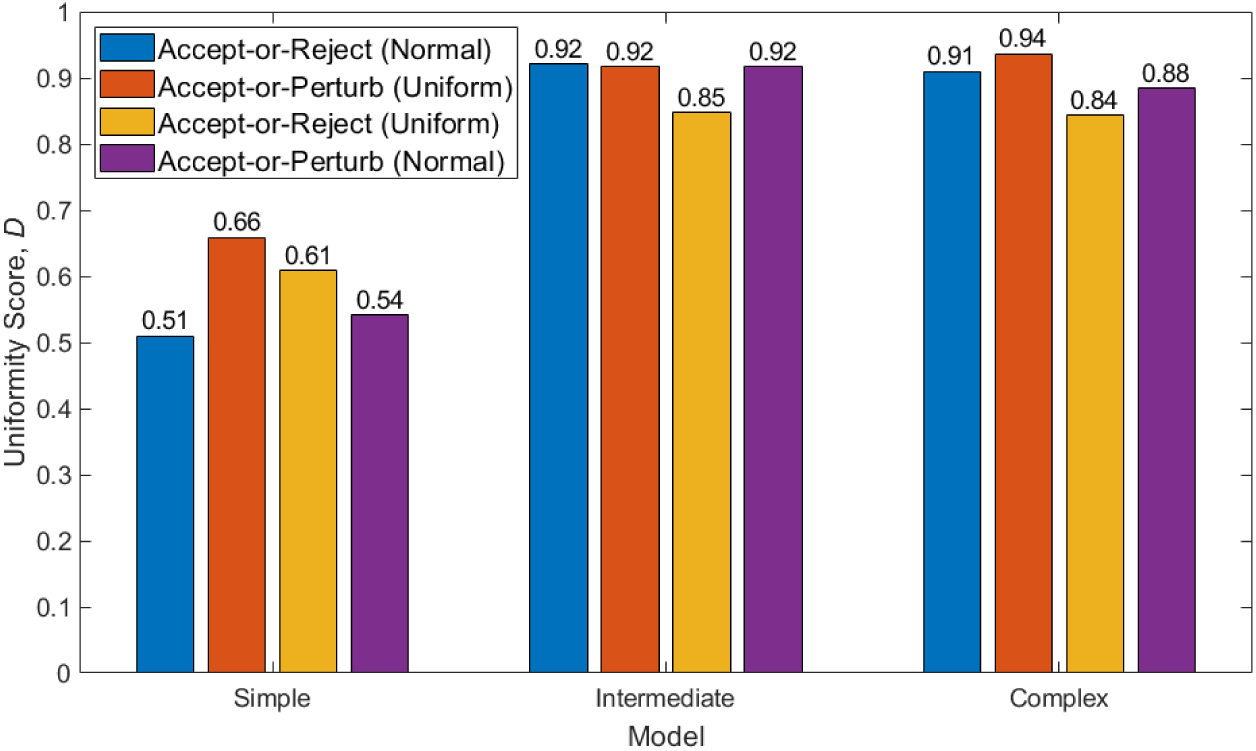
Extent to which plausible population spans the feasible trajectory space as measured using the uniformity score, *D*, at Day 16. Results are shown across all three models using the standard pairing of priors and inclusion/exclusion criteria (accept-or-reject with a normal prior and accept-or-perturb with a uniform prior) and for the “flipped” cases (accept-or-reject with a uniform prior and accept-or-perturb with a normal prior).

**Fig. S5.**
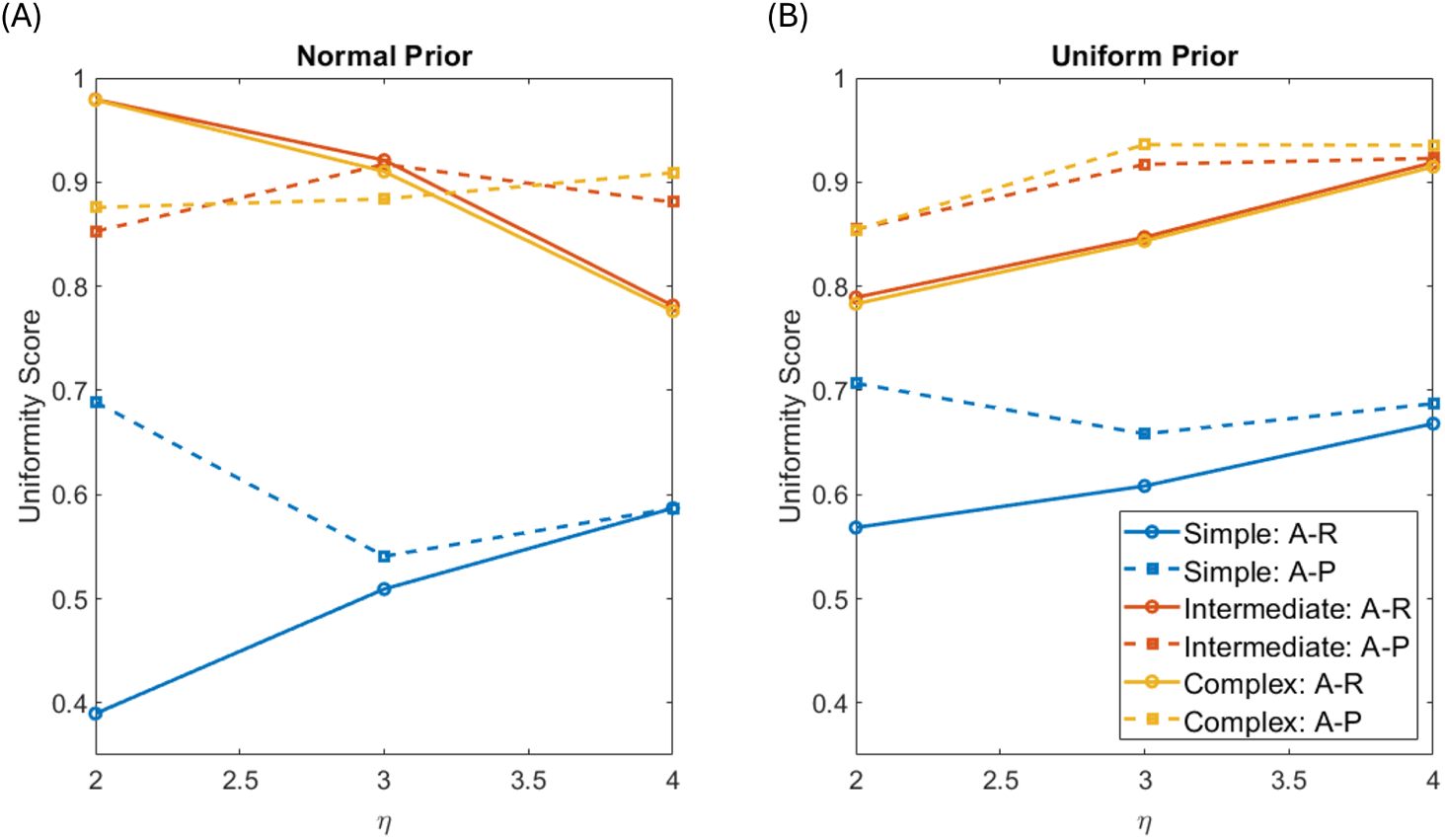
Extent to which plausible population spans the feasible trajectory space as measured using the uniformity score, *D*, at Day 16. Results are shown as a function of the extent of the feasible region, *η*, across all three models and inclusion/exclusion criteria. (A) Using a normal prior. (B) Using a uniform prior. Here, A-R stands for accept-or-reject, and A-P stands for accept-or-perturb. We observe that the uniformity score is more variable across *η* when accept-or-reject is used (solid curves) compared to when accept-or-perturb is used (dashed curves).

**Fig. S6.**
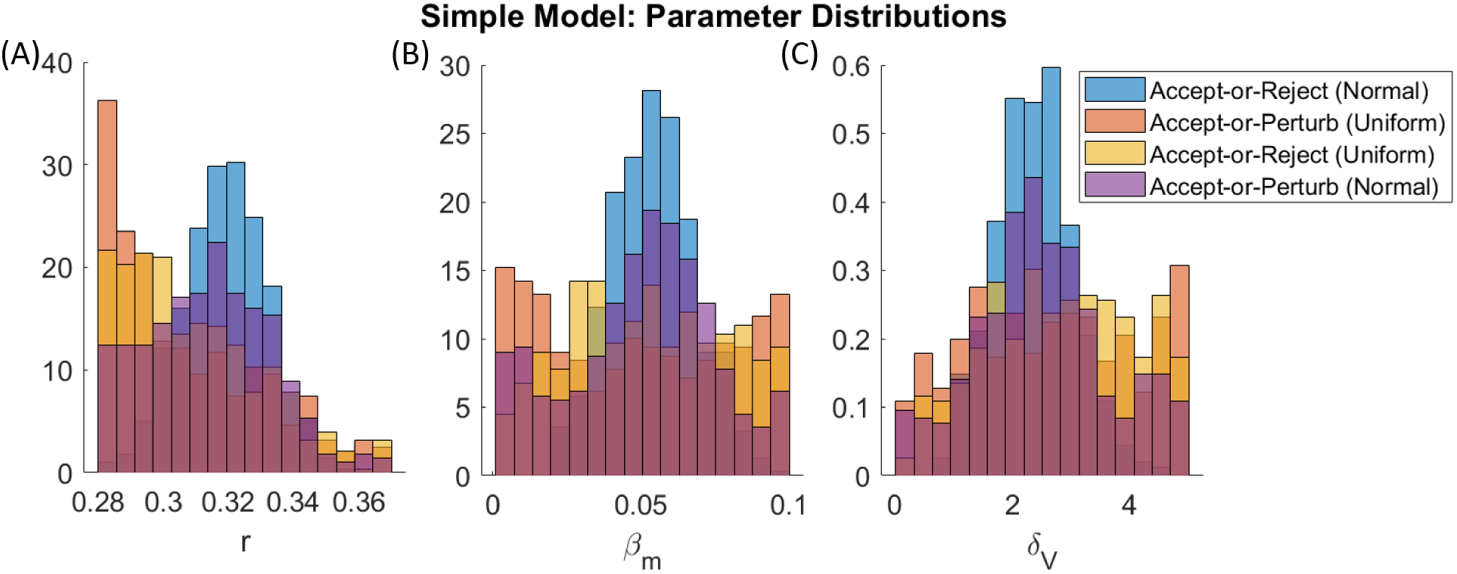
Posterior distributions for the parameters included in the plausible population using the simple model. (A) Tumor growth rate, *r*. (B) Viral infection rate, *β_m_*. (C) Viral decay rate, *δ_V_* . Results are shown for each inclusion/exclusion criterion, using both a normal and uniform prior distribution.

**Fig. S7.**
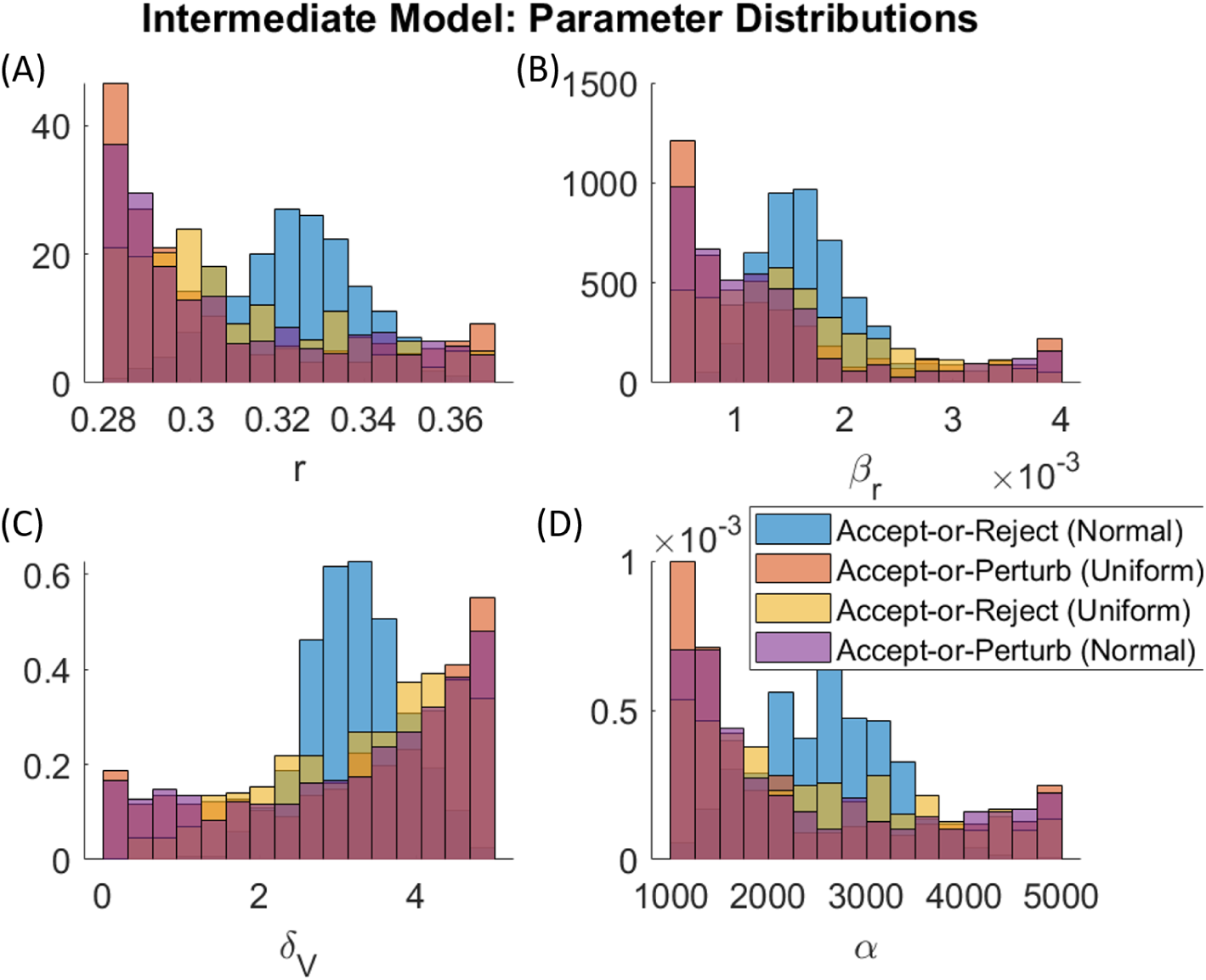
Posterior distributions for the parameters included in the plausible population using the intermediate model. (A) Tumor growth rate, *r*. (B) Viral infection rate, *β_r_*. (C) Viral decay rate, *δ_V_* . (D) Viral burst size, *α*. Results are shown for each inclusion/exclusion criterion, using both a normal and uniform prior distribution.

**Fig. S8.**
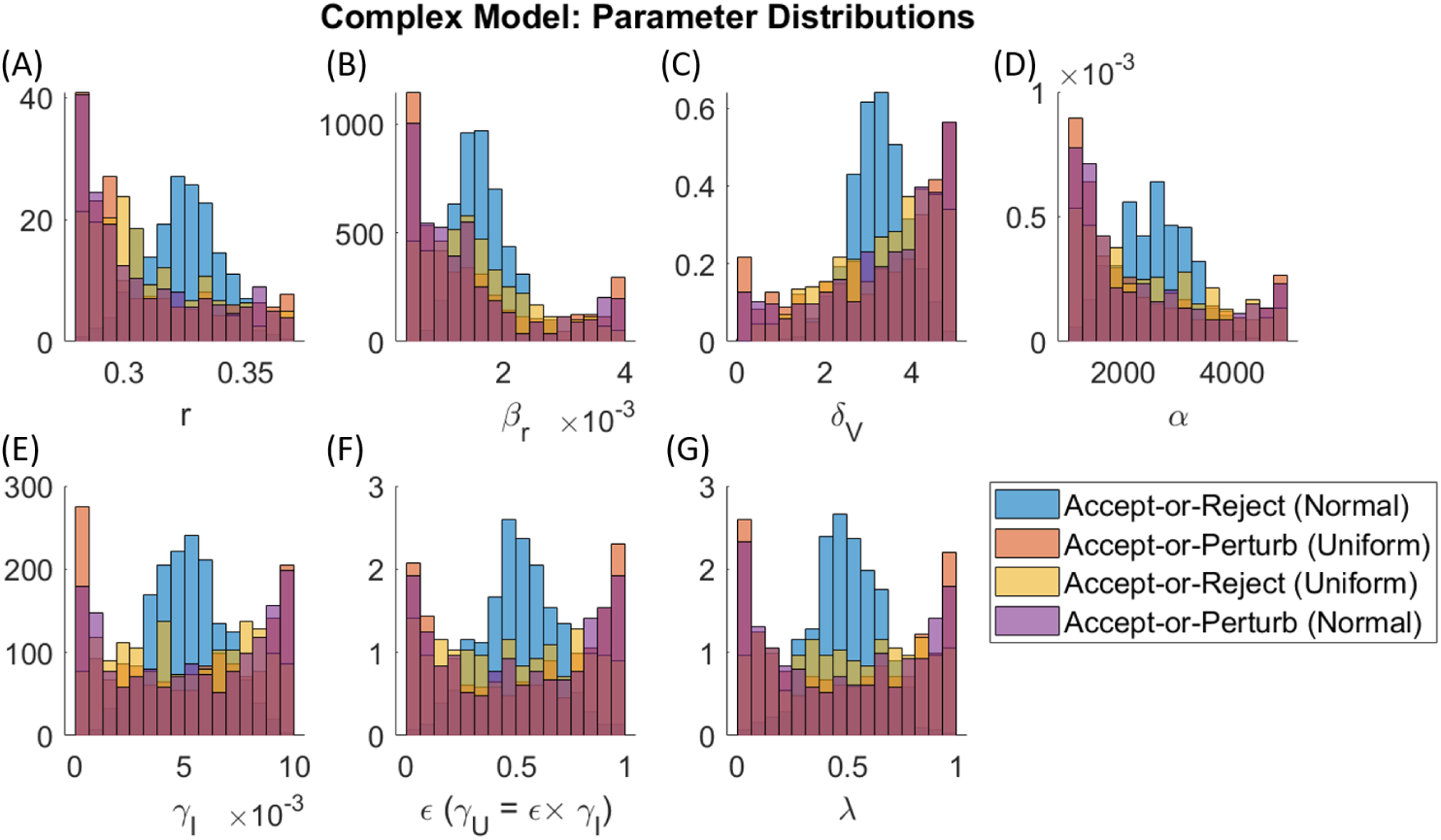
Posterior distributions for the parameters included in the plausible population using the complex model. (A) Tumor growth rate, *r*. (B) Viral infection rate, *β_r_*. (C) Viral decay rate, *δ_V_* . (D) Viral burst size, *α*. (E) Immune-kill rate of infected cell, *δ_I_* . (F) Scaling factor for immune-kill rate of uninfected cells, *ɛ* where *δ_U_* = *ɛδ_I_* . (G) Immune cell recruitment rate, *λ*. Results are shown for each inclusion/exclusion criterion, using both a normal and uniform prior distribution.

**Fig. S9.**
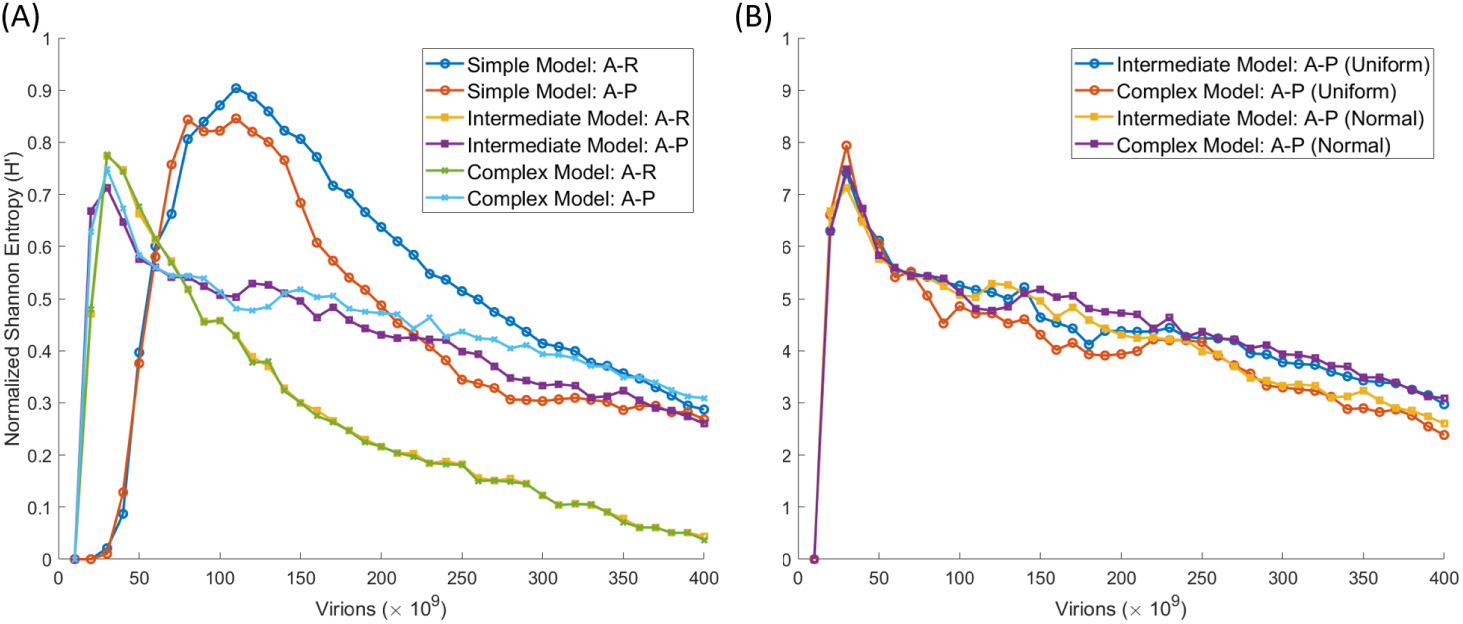
Normalized Shannon entropy of the virtual clinical trial outcomes for (A) the simple, intermediate and complex models using accept-or-reject with a uniform prior, and accept-or-perturb with a normal prior. (B) Only the intermediate and complex model using accept-or-perturb with either a normal or uniform prior.

**Fig. S10.**
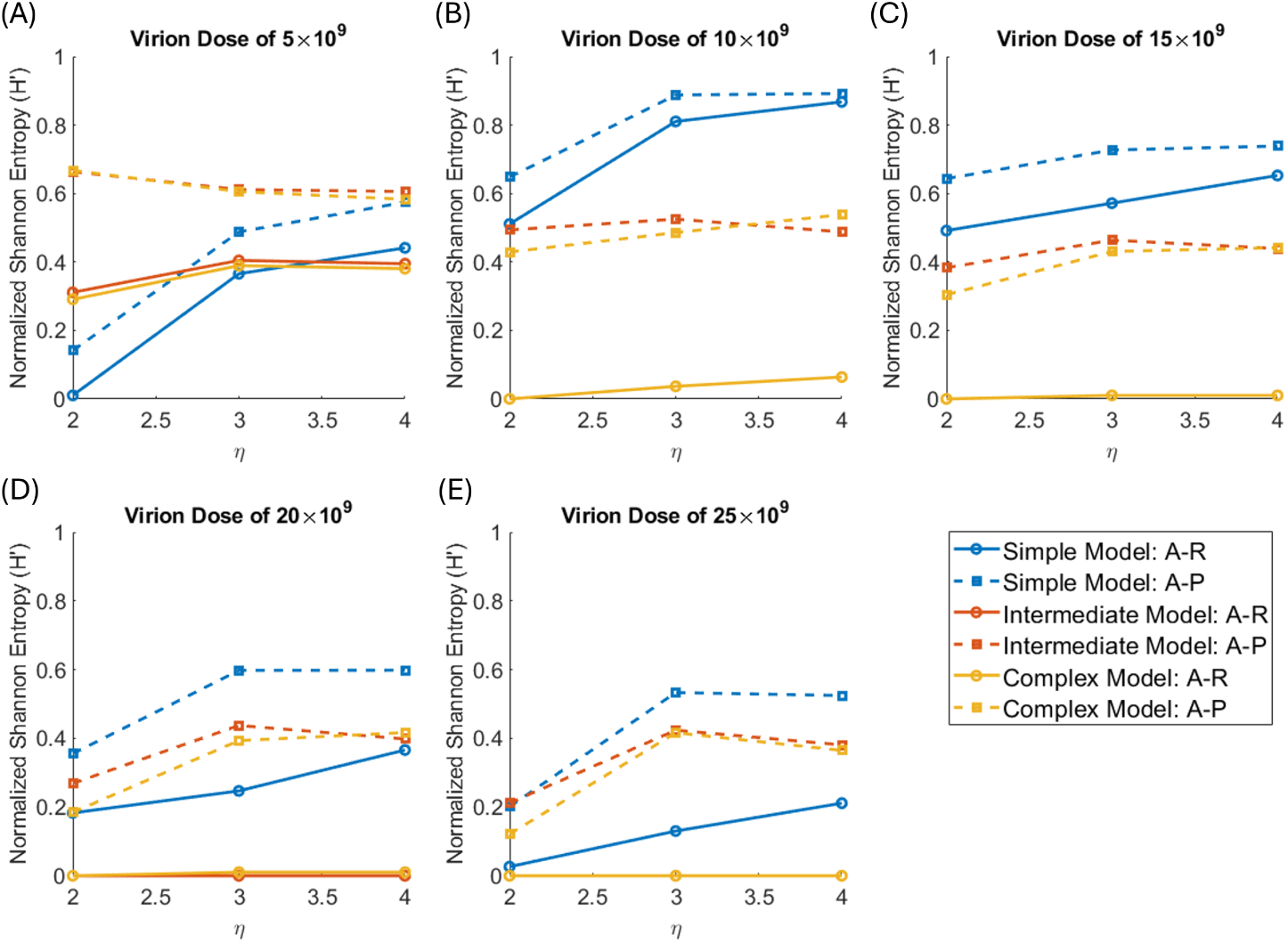
Normalized Shannon entropy of the virtual clinical trial outcomes as a function of the extent of the feasible region, *η*. Results are shown for the simple, intermediate and complex models using the standard pairing of accept-or-reject with a normal prior, and accept-or-perturb with a uniform prior. Virion doses considered are (A) 5 *×* 10^9^, (B) 10 *×* 10^9^, (C) 15 *×* 10^9^, (D) 20 *×* 10^9^, and (E) 25 *×* 10^9^. Across nearly all scenarios, *η* = 2 yields the highest homogeneity (lowest *H^′^*) in VCT outcomes.

